# Integrative multi-ancestry genetic analysis of gene regulation in coronary arteries prioritizes disease risk loci

**DOI:** 10.1101/2023.02.09.23285622

**Authors:** Chani J. Hodonsky, Adam W. Turner, Mohammad Daud Khan, Nelson B. Barrientos, Ruben Methorst, Lijiang Ma, Nicolas G. Lopez, Jose Verdezoto Mosquera, Gaëlle Auguste, Emily Farber, Wei Feng Ma, Doris Wong, Suna Onengut-Gumuscu, Maryam Kavousi, Patricia A. Peyser, Sander W. van der Laan, Nicholas J. Leeper, Jason C. Kovacic, Johan L.M. Björkegren, Clint L. Miller

## Abstract

Genome-wide association studies (GWAS) have identified hundreds of genetic risk loci for coronary artery disease (CAD). However, non-European populations are underrepresented in GWAS and the causal gene-regulatory mechanisms of these risk loci during atherosclerosis remain unclear. We incorporated local ancestry and haplotype information to identify quantitative trait loci (QTL) for gene expression and splicing in coronary arteries obtained from 138 ancestrally diverse Americans. Of 2,132 eQTL-associated genes (eGenes), 47% were previously unreported in coronary arteries and 19% exhibited cell-type-specific expression. Colocalization analysis with GWAS identified subgroups of eGenes unique to CAD and blood pressure. Fine-mapping highlighted additional eGenes of interest, including *TBX20* and *IL5*. Splicing (s)QTLs for 1,690 genes were also identified, among which *TOR1AIP1* and *ULK3* sQTLs demonstrated the importance of evaluating splicing events to accurately identify disease-relevant gene expression. Our work provides the first human coronary artery eQTL resource from a patient sample and exemplifies the necessity of diverse study populations and multi-omic approaches to characterize gene regulation in critical disease processes.

**Study Design Overview:** 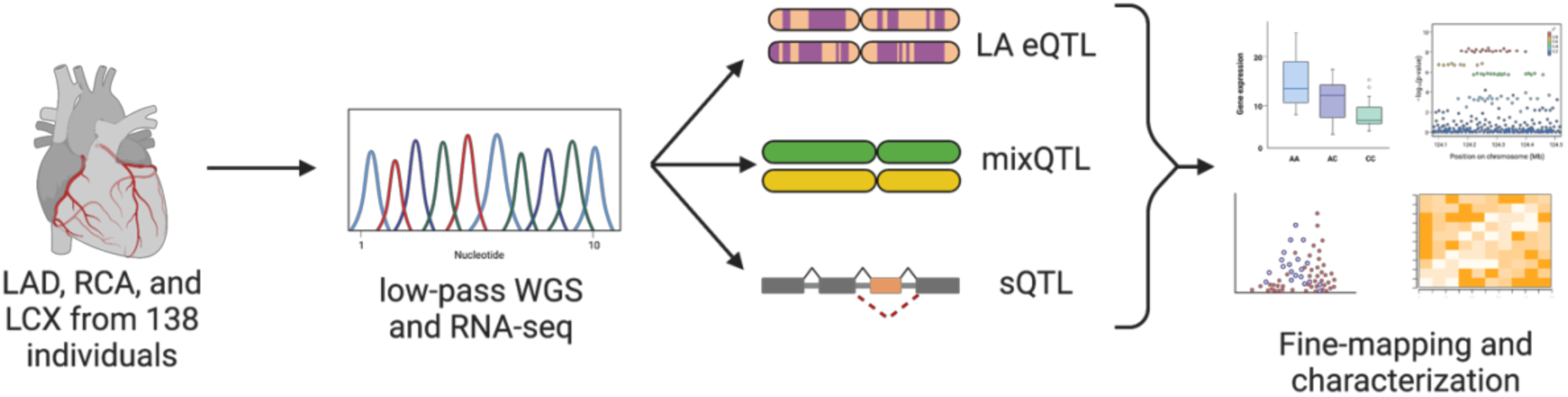

## Introduction

Coronary artery disease (CAD) is the leading cause of death worldwide, and results from chronic inflammatory processes involving both genetic and environmental risk factors. CAD manifests as the development of atherosclerotic plaques in the coronary arteries of the heart, which can lead to erosion or plaque rupture, and ultimately myocardial infarction. Genome-wide association studies (GWAS) have now reported more than 400 independent loci for CAD and related clinical outcomes.^1–11^ As with other common complex traits, the majority of lead CAD GWAS variants reside in non-coding genomic regions, implicating regulatory effects on gene expression.^12^ Previous studies have mapped CAD GWAS variants to specific cell types in the vessel wall (e.g., smooth muscle cells (SMC),^13^ endothelial cells, and immune cells)^14^ and refined candidate c*is*-acting regulatory elements (e.g., promoters, enhancers) responsible for context-specific gene expression patterns.^14,15^ However, cultured vascular cells do not fully recapitulate their cell phenotype *in vivo*: for instance, over multiple passages SMCs reprogram toward a fibroblast-like state accompanied by rapid loss of differentiated marker gene expression.

Fine-mapping GWAS loci can help prioritize candidate causal variants within association signals, but identifying the causal variant or target gene within a locus with many associated variants can still be difficult. Overrepresentation of European- and East Asian-ancestry populations in most GWAS to date has also limited both the capacity to identify independent associations within a locus and the generalizability of findings to global populations.^16–18^ Furthermore, genes within most CAD loci have not been associated with traditional risk factors (e.g., lipid levels or cholesterol metabolism), suggesting molecular mechanisms underlying physiological effects on the coronary artery vessel wall itself.

Molecular quantitative trait locus mapping in a disease-relevant tissue or cell line is a powerful approach to prioritize candidate causal genes and underlying mechanisms for complex GWAS loci.^19^ Prior studies have identified CAD-relevant expression QTL (eQTL) in bulk arterial tissues^20^ or specific vessel wall cell types, including human aortic endothelial cells,^21^ human coronary artery smooth muscle cells (HCASMC),^13,22^ and monocytes.^23^ Similar to CAD GWAS and other eQTL studies, published summary statistics represent exclusively or primarily European-ancestry populations, and often lack detailed phenotyping or clinical information on the patients/participants.

To identify variants causally associated with coronary artery-specific gene expression and facilitate fine-mapping of CAD GWAS associations, we performed an eQTL mapping study in coronary artery tissues from an ancestrally diverse American patient-derived sample. We utilized two different methods to identify expression and splicing QTLs in human coronary artery tissue, followed by bioinformatic characterization of potential eGene-phenotype associations. Our results not only highlight new coronary artery eQTLs at promising GWAS loci such as *TBX20* but also replicate and refine eQTLs previously reported in other arterial tissues, including *ARHGAP42*. This work improves the capacity to characterize gene regulation in coronary artery tissue through all stages of CAD progression. This coronary artery eQTL dataset will therefore be a highly beneficial resource for better characterization of functional variants and molecular mechanisms driving CAD development.

## Results

### Study overview for transcriptomic profiling of human coronary artery

We conducted transcriptome-wide QTL mapping of autosomal gene expression in human coronary artery tissue samples from explanted transplant tissue as well as samples collected from rejected transplant donors (**Materials and Methods, Table S1**). An overview of our sample characteristics, and primary diagnoses are described in **Figure 1** and **Tables S1-3.** The study comprised 138 individuals from 19 to 72 years old, with 30% being female. While 57% of these individuals were of exclusively European ancestry, 15% were of majority South Asian ancestry, 5% East Asian, 5% Indigenous American, 7% African and 10% were majority of admixed genetic ancestry, representing the broad genetic diversity of the American population (**Figures 1a-c, S1**). Samples were derived from all three major coronary arteries (i.e., left anterior descending coronary artery [LAD], right coronary artery [RCA], and left circumflex artery [LCX], **Figure 1d**), and explanted samples represented various patient diagnoses (**Figure 1e, Table S1**). Up to 5.83 million variants genotyped via low-pass whole-genome sequencing were included depending on method-specific allele frequency and annotation requirements (described in Methods, **Table S2**).

**Fig 1.**
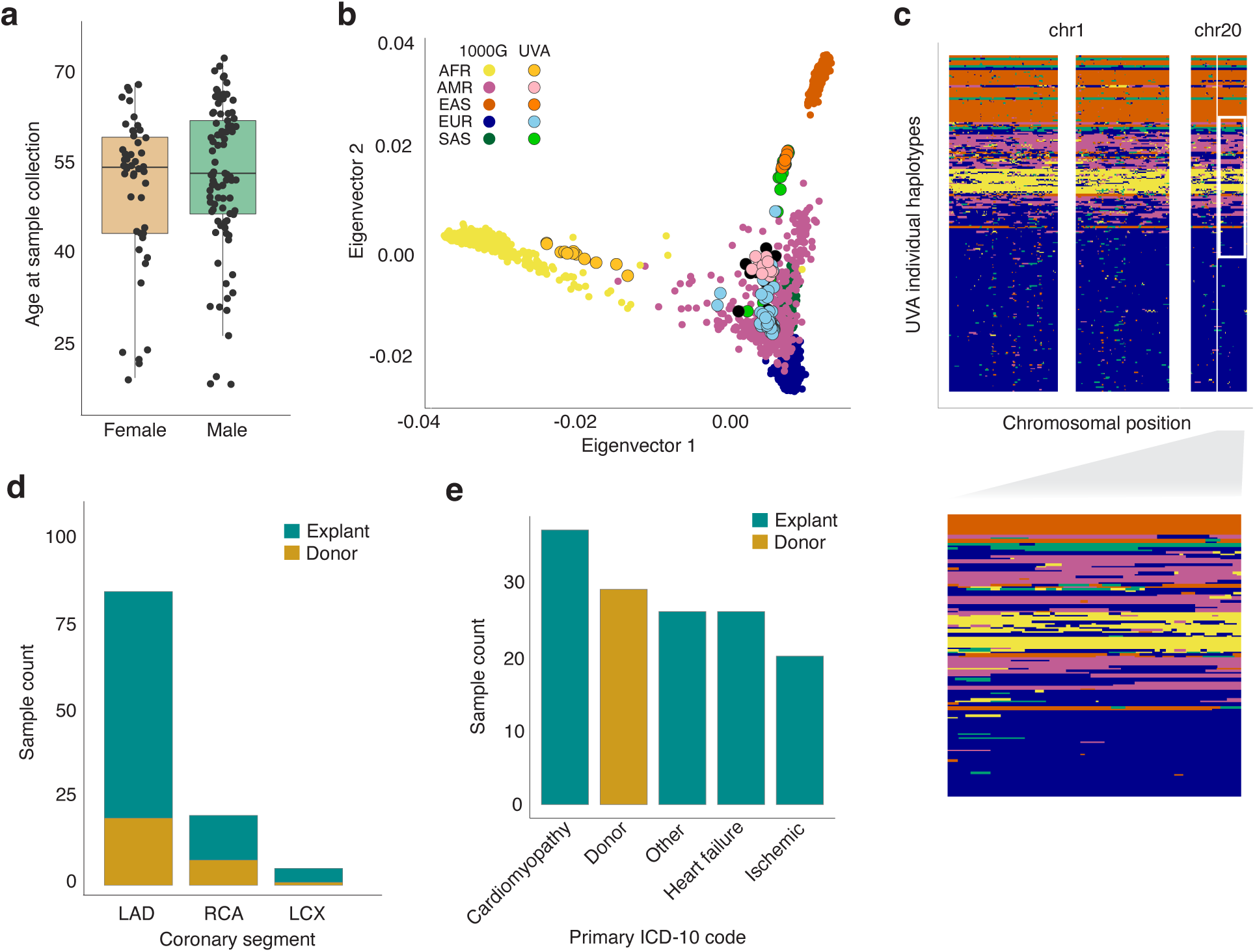
Overview of patient and sample characteristics. (**a**) Boxplot showing the age range of study participants (y-axis) in females and males (x-axis; orange and green, respectively). (**b**) Genetic ancestry principal components 1 (x-axis) and 2 (y-axis) mapped onto the 1000 Genomes Phase 3 reference population (bold color dots), with color corresponding to Gencove-assigned majority ancestry in our samples (lighter color dots with black outline). (**c**) Local ancestry inference reveals a complex genetic substructure for individuals with ancestral admixture, with each row in the plot representing inferred local ancestry for one haplotype of one study participant and x-axis representing position on respective chromosomes. Inset, zoomed in region on chr 20 showing genetic substructure for a subset of the individual haplotypes. (**d**) Number (y-axis) of coronary artery segments by type (x-axis) used for RNA isolation in samples from explants (turquoise) and rejected donors (gold). (**e**). Number (y-axis) of study population members according to primary ICD 10 diagnosis for explants (turquoise) or rejected donor status(gold, x-axis).

We performed total RNA sequencing to a median depth of 102.6 million reads per sample (**Table S3**), to profile both protein-coding and non-coding RNAs (**Figure S1**). To determine the similarity of our expression profiles to bulk RNA profiles of other tissues in GTEx as well as cultured human coronary artery smooth muscle cells (HCASMC), we performed multidimensional scaling. Our samples form a cluster located near the left ventricle, muscle, pancreas, fibroblasts, and liver tissues (**Figure S1**). This distinct but proximal clustering aligns with expectations given differences in sample collection/storage methods and cold ischemia times (time lapsed after cessation of blood flow). Since eQTL studies have primarily been performed in genetically homogeneous populations, information on preferred methods for inclusive study populations is limited. We therefore applied two complementary approaches to attempt to maximize power for identifying associations that may not be globally present, i.e., by evaluating haplotype-specific associations (“mixQTL” for the total population or “mixQTL_EUR_” for the 100% European ancestry subset analysis) or ancestry-specific associations (local ancestry-adjusted, henceforth referred to as “LA”).^24,25^ After filtering for method-specific criteria, up to 20,100 autosomal protein-coding genes and lncRNAs met inclusion criteria for mixQTL and local ancestry eQTL and sQTL analysis.

### Coronary artery eQTL discovery

To identify genetic variants associated with gene expression in our diverse coronary artery tissue cohort, we performed eQTL analyses incorporating haplotype-specific (mixQTL)^24^ or local ancestry (LA) information. Overall, we identified 2,132 and 793 coronary artery eGenes using mixQTL or LA, respectively (**Tables 1, S4, S5, Figure 2a**). Between LA and mixQTL analyses, 457 shared eGenes were identified (**Figure 2b**); 45 lead SNPs were common to both approaches (**Table S6**). Of note, across both methods we report 561 total discovery eGenes (351 mixQTL, 163 LA, 210 mixQTL_EUR_; 200 protein-coding genes, 361 lncRNAs) with no expression QTLs reported in any arterial tissue in GTEx or STARNET, including genes with established roles in vascular cell types (e.g., lipase G, endothelial type [*LIPG*] and AKT Serine/Threonine Kinase 3 [*AKT3*]).^26,27^ Forty percent of discovery eQTLs were >100kb from the gene transcription start site, which is in line with long-standing evidence of both short- and long-range cis-acting regulatory mechanisms.^28,29^ We report mixQTL results from the entire study population as our primary findings given the higher statistical power of this method.

**Table 1.**
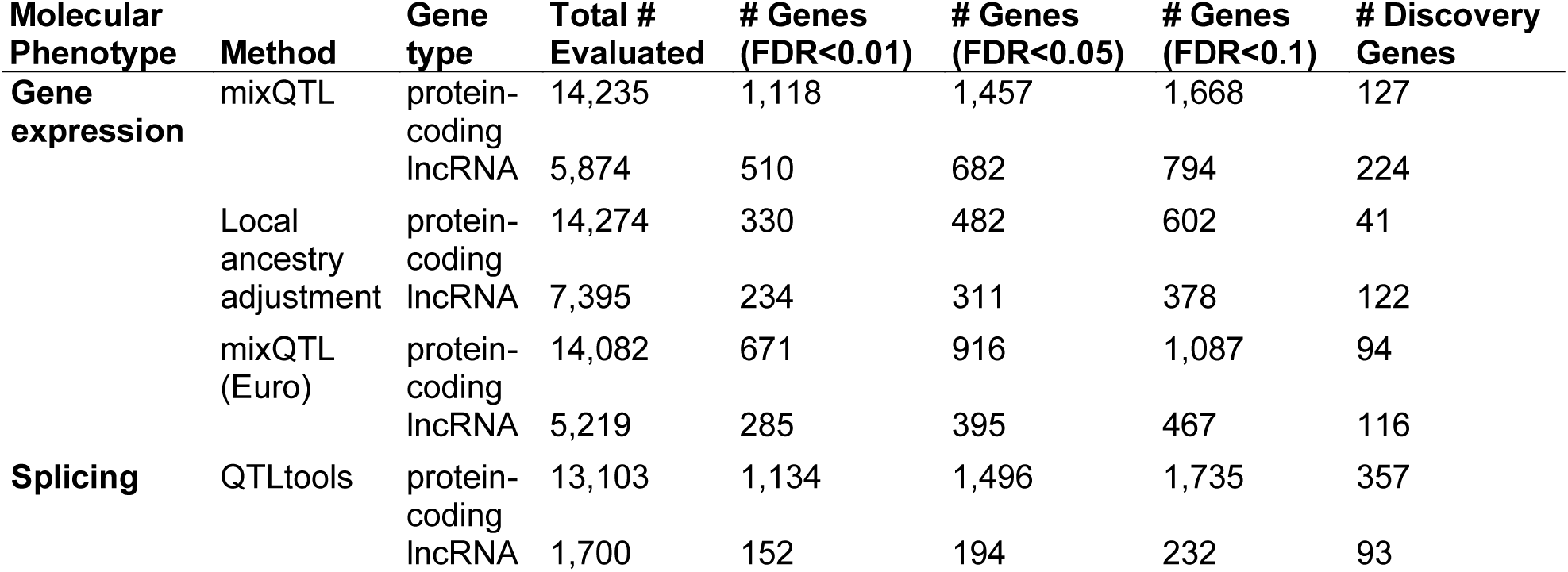
Discovery expression and splicing QTLs in human coronary artery tissue.

| **Molecular Phenotype** | **Method** | **Gene type** | **Total # Evaluated** | **# Genes (FDR<0.01)** | **# Genes (FDR<0.05)** | **# Genes (FDR<0.1)** | **# Discovery Genes** |
| --- | --- | --- | --- | --- | --- | --- | --- |
| **Gene expression** | mixQTL | protein-coding | 14,235 | 1,118 | 1,457 | 1,668 | 127 |
|  |  | lncRNA | 5,874 | 510 | 682 | 794 | 224 |
|  | Local ancestry adjustment | protein-coding | 14,274 | 330 | 482 | 602 | 41 |
|  |  | lncRNA | 7,395 | 234 | 311 | 378 | 122 |
|  | mixQTL (Euro) | protein-coding | 14,082 | 671 | 916 | 1,087 | 94 |
|  |  | lncRNA | 5,219 | 285 | 395 | 467 | 116 |
| **Splicing** | QTLtools | protein-coding | 13,103 | 1,134 | 1,496 | 1,735 | 357 |
|  |  | lncRNA | 1,700 | 152 | 194 | 232 | 93 |

**Fig 2.**
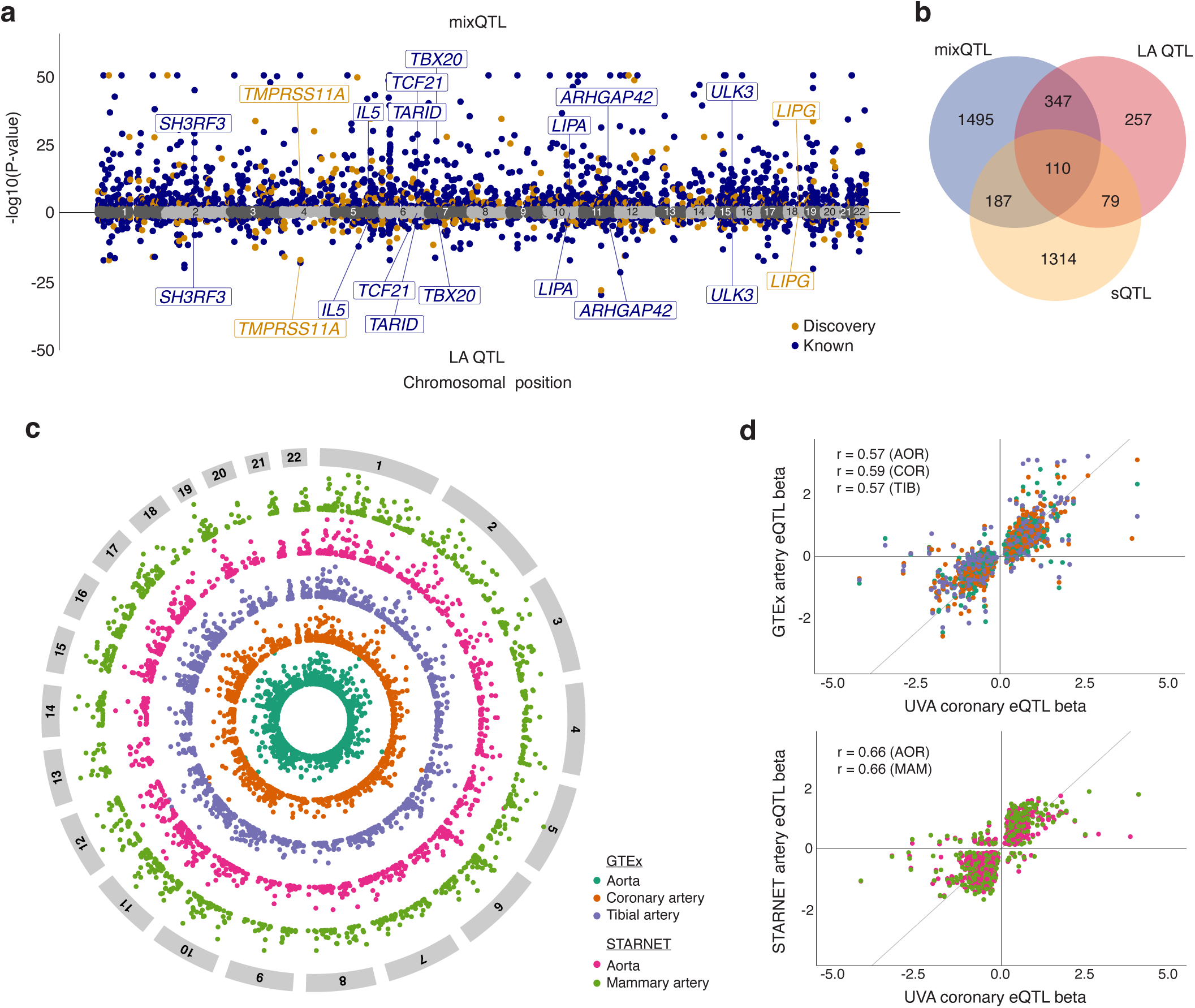
Overview of eQTL analysis and generalization to published arterial eQTLs. (**a**) Miami plot of lead eQTLs for mixQTL (top) and local ancestry (LA)-adjusted (bottom). Navy blue and orange dots represent reported and discovery eGenes (pBH<0.05), respectively; gray dots represent non-significant genes. A subset of top eGenes are labeled for clarity. (**b**) Venn diagram showing the overlap of mixQTL and LA based eGenes and Leafcutter sQTL sGenes. (**c**) Circos plot portraying generalized (UVA pBH<0.05) published arterial eGenes from GTEx AOR (blue green), COR (orange), and TIB (purple), and STARNET AOR (pink) and MAM (light green) tissues, with significance increasing toward the outer edge of the circle. (**d**) Direction of effect for genes in which the UVA lead eQTL was significant (pBH<0.05) in any of the aforementioned tissues using the same color scheme for GTEx (top) and STARNET (bottom). Pearson’s r correlation coefficients shown for overlapping significant UVA coronary eQTL detected in GTEx or STARNET eQTL with tissue indicated in parentheses. AOR: aorta; COR: coronary artery; TIB: tibial artery; MAM: mammary artery.

### Overview of mixQTL results

Of the 2,132 protein-coding or lncRNA mixQTL eGenes, 16% (n=351) have not been previously reported in published arterial tissue QTL studies (**Table S4**). In concordance with published QTL studies, most eQTLs were annotated as intronic to their eGenes (2,232 of 2,878 total annotations for 746 eGenes (**Figure S2**)). Only two percent of lead eQTLs (n=59) were protein-coding within their respective eGene, implicating the regulation of gene expression through transcriptional, splicing, or epigenetic mechanisms.^20^ Thirty-nine and 119 eGenes were identified based solely on allele-specific expression and total read count tests, respectively. Overall, 1,779 published arterial eGenes had significant eQTLs in our study sample (**Figure 2c**, **Table S7**). Fewer than 5% of shared associations had the same lead eQTL, but among shared lead eQTLs we observed 98% directional consistency (64% of all replicated lead eQTLs, **Figure 2d, Figure S2**). Nearly one in five eGenes exhibited cell-type-specific expression in a coronary artery single-cell RNA sequencing reference dataset (cell-specificity expression score ≥0.7, **Figure S2, Table S8**),^30^ and functional enrichment analysis revealed several pathways for cell adhesion and inflammation (**Table S9**).^31^

### Local-ancestry-adjusted eQTLs

With regard to local-ancestry-adjusted analyses, 337 eGenes were identified that did not exceed a FDR of 5% in the overall mixQTL analysis, demonstrating the merit of incorporating multiple approaches in a diverse study sample with genetic admixture (**Table S5**). Among LA-specific eGenes was YY1-associated protein 1 (*YY1AP1*), which has no reported coronary artery eQTLs but has been associated with vascular diseases including Grange Syndrome and sudden coronary artery dissection.^32,33^ Seventeen LA lead variants were monomorphic in the 1000G EAS superpopulation (**Table S10A**). Despite high-confidence calling of 1000G continental ancestries in our study sample, the small numbers of shared haplotypes at any given locus likely limited our ability to identify ancestry-specific associations using this method. Using mixQTL, 54 eGenes with lead SNPs monomorphic in one or more 1000G superpopulations were identified; four overlapped with the LA results and lead eQTLs were either shared or in high linkage disequilibrium (LD) (**Tables S10B, S11**). In combination, these results indicate both improved power for mixQTL compared to a traditional unphased approach as well as the benefit of including genetically diverse individuals in identifying associations that improve effective sample size across the lower end of the global allele frequency range.

### Colocalization of eQTLs

We next evaluated the overlap between coronary artery gene expression and genetic associations with CAD and intermediate risk factors including blood pressure, cholesterol, and arterial calcification traits. Across all phenotypes, 108 GWAS association signals colocalized with eQTLs, including 25 discovery eGenes (**Figure 3a**, **Table S12**). Thirty-one eGenes exhibiting cell type specificity colocalized to one or more GWAS, including Rho GTPase-Activating Protein 42 (*ARHGAP42*) in pericytes and discovery eGenes Lipase G (*LIPG)* and Adhesion G Protein-Coupled Receptor G6 (*ADGRG6*) in endothelial cells (**Table S8**). We further evaluated eQTLs using summarized Mendelian randomization (SMR),^34^ which had fewer associations but notable overlap with colocalization (25 overlapping signals and 18 unique to SMR, **Figure S2, Table S13**).

**Fig 3.**
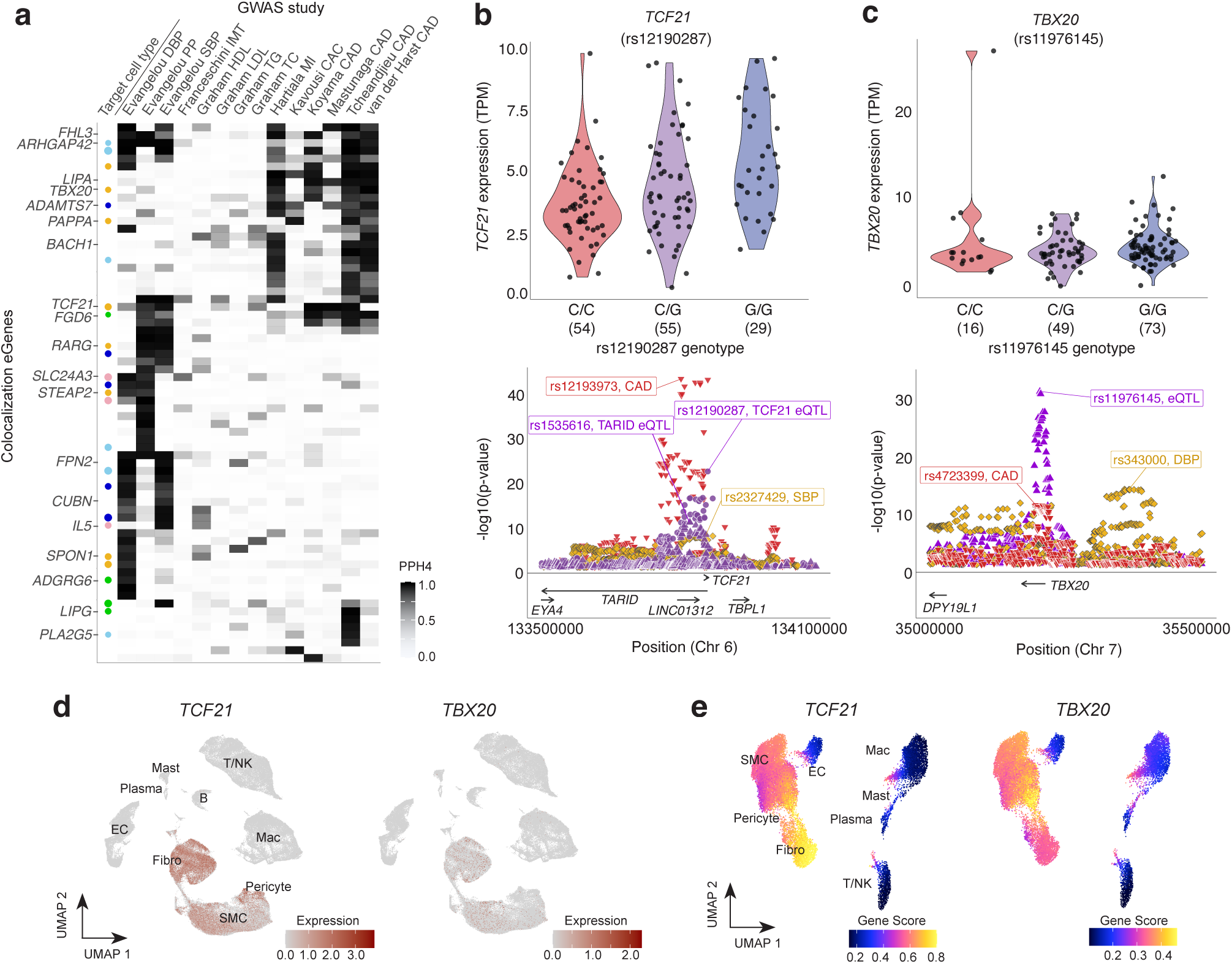
Colocalization reveals trait- and cell-type-specific associations. (**a**) GWAS colocalization to eGene associations: each column represents the -log10(p-value) of the study (first author last name) and relevant GWAS trait, with intensity of shading corresponding to a higher posterior probability of a shared association at that locus. Each row represents one protein-coding gene with a PPH4≥0.8 in at least one GWAS. The leftmost column (“Target cell type”) indicates cell-type specificity for SMC (dark blue), pericyte (light blue), endothelial (green), fibroblast (gold), or blood (pink) cells. Size of the circle represents the CELLEX combined gene score (range: 0.7 to 1.0). (**b,c**). Top: Normalized expression (y-axis) by lead eQTL genotype (x-axis) for *TCF21* (**b**) and *TBX20* (**c**). Bottom: Regional association plots showing overlap between our study (violet) and GWAS associations for CAD (red triangle) and blood pressure traits (gold diamonds). (**d,e**). Human artery atherosclerosis single-cell RNA sequencing (left) and single-nuclear ATAC sequencing (right) showing *TCF21* and *TBX20* gene expression and chromatin accessibility based gene scores, respectively, with changes indicated by intensity of red (left) and pink/yellow (right).

We observed strong evidence of colocalization and generalization at the *TCF21*/*TARID* locus. *TCF21*, a known regulator of the SMC phenotype transition to fibromyocytes in plaque,^35,36^ exhibited a strong eQTL for the same variant, rs12190287, in our study as well as in STARNET aorta (AOR) and GTEx coronary artery (COR) samples (**Figures 3b, S3**). The *TCF21* eQTL overlapped with the association for the adjacent lncRNA *TARID* (TCF21 antisense RNA inducing promoter demethylation). Our lead *TARID* eQTL, rs1535616, was also significant in GTEx COR, though not for other arterial tissues (**Figure S3**). Interestingly, GTEx COR exhibited similarly strong associations at both ends of the *TARID* coding region, while AOR showed a much stronger association at the 3’ end (**Figure S3**). *TCF21* and *TARID* also colocalized with CAD and BP trait associations but not coronary artery calcification or cholesterol traits (**Figure 3b**), suggesting the mechanism for this known CAD locus may be functioning via a causal blood pressure pathway. Coronary artery single-cell RNA and Assay for Transposase-Accessible Chromatin sequencing (scRNA-seq, snATAC-seq) reference datasets show that *TARID* and *TCF21* are both expressed more highly and more accessible in fibroblasts and SMCs (including SMC-derived fibromyocytes resulting from SMC phenotypic switching during atherosclerosis, **Figures 3d,e, S3**).^30,37^

*TBX20*, coding for transcription factor T-box 20, is another gene with established cardiac development and disease associations,^38^ however the mechanism of its genetic regulation or link to CAD risk remains unclear. Our lead eQTL, rs11976145 (**Figure 3c**), is located in the fourth intron and colocalizes with a CAD GWAS signal but is independent from the DBP association at the same locus, suggesting a complex approach may be required to ascertain disease-relevant mechanisms underlying transcriptional regulation. *TBX20* is most abundant in cardiac muscle and vascular tissues in GTEx, with coronary artery reference expression predominantly restricted to SMCs and fibroblasts (**Figure 3d,e**). While *TBX20* has no reported eQTLs for GTEx arterial tissues, we identify an overlapping association between the eQTL for STARNET (AOR), for which the lead eQTL rs10249005 is in high LD (r^2^≥0.94) in all European-ancestry 1000 Genomes reference populations (**Table S11**).

### Fine-mapping discovery eQTLs

Next, we used a combination of methods to identify both credible sets (CS) and independent associations within previously unreported coronary artery eGenes. First, we used the Bayesian mixFine function from the mixQTL package to identify independent credible sets for significant associations (**Table S14**). Only 3% of eGenes with convergine association signals fine-mapped to a single variant (n=44 of 1,388). In total, 91% of CS contained the lead eQTL, which was the sole credible variant for 29 eGenes. Eighty-three eGenes exhibited multiple independent eQTL signals.

We additionally employed FastPaintor to fine-map associations with epigenomic annotations and relevant GWAS. 1,714 eGenes had a sufficient number of eQTLs to converge for fine-mapping annotation using either BP or CAD GWAS associations. Across both traits and three epigenetic marks, CS for 78 eGenes were narrowed to a single likely causal variant, and 11% contained the lead eQTL (**Table S15A,B**). Regardless of annotation combination, most CS (>75%) were narrowed to five or fewer SNPs (**Figure S4a**). While the majority of fine-mapped associations comprised CS containing the same variants regardless of epigenetic annotation, they often differed by GWAS annotation. The 187 loci for which at least one variant was shared between BP trait- and CAD-annotation exhibited lower p-values and higher proximity of lead eQTL to the eGene TSS (mean distance 25.2kb, **Figure S4b**).

We demonstrate the utility of combining multi-omic data with fine-mapped associations in *ARHGAP42*, a Rho-A GTPase activating protein with functional evidence for disease relevance, and *IL5*, a discovery coronary eGene. Lead *ARHGAP42* eQTL rs2455569 exhibited similar effect direction and magnitude in STARNET aorta tissues (**Figure S5a,b**), and decreased *ARHGAP42* expression in AOR was significantly associated with case status in STARNET (**Table S16**). The *ARHGAP42* eQTL overlapped with GTEx AOR and COR associations, and colocalized with both BP traits and CAD GWAS associations (**Table S12**, **Figure S5c,d**). ARHGAP42 (also known as GRAF3) regulates vascular tone via expression predominantly in mural cells and fibroblasts (**Figure S5e,f**),^30,37,39^ and insufficiency causes hypertension.^40,41^ rs2455569 is located in a pericyte-specific chromatin-accessibility peak in the first intron of *ARHGAP42*, with fine-mapping highlighting proximal SNPs encompassing a region accessible in multiple vascular cell types (**Figure 4a-c**).^37^ Interestingly, rs604723, an eQTL 25kb upstream in high LD with rs2455569 (*p*_adj_=4.6E-13, **Table S11**), has been shown to modulate *ARHGAP42* expression in SMCs via SRF binding.^42^ Rs604723 is also predicted to disrupt a binding motif for STAT6, which has been implicated in the proliferation of vascular SMCs in an injury-response murine model,^43^ suggesting multiple avenues for the effects of this locus on *ARHGAP42* and its downstream effects on disease processes (**Figure 4d**).

**Fig 4.**
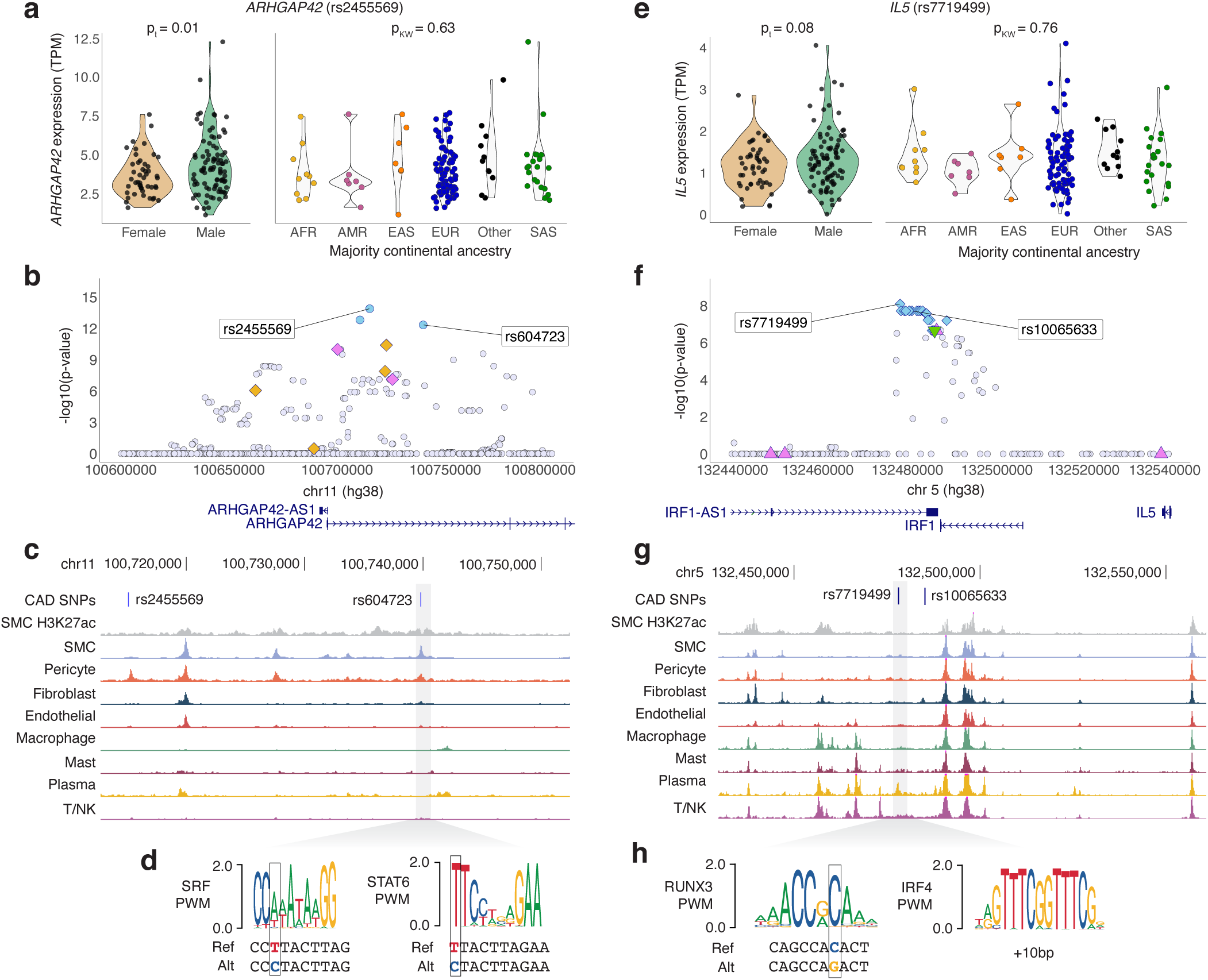
Fine-mapping identifies candidate causal variants for *ARHGAP42* and *IL5* eQTLs at known GWAS loci. (**a**) *ARHGAP42* association driven by lead eQTL rs2455569, exhibiting differences in expression by genotype but not sex or majority continental ancestry. (**b**) Regional association plot depicting variants in PAINTOR credible sets specific to BP (violet) or CAD (golden) GWAS annotations, mixFine only (light blue), and variants not in any credible set (light gray). (**c**) Variants of interest (rs604723 and rs2455569) are indicated by blue lines and rs604723 is highlighted in gray box, and corresponds to UCSC Genome Browser tracks indicating cell-type-specific chromatin accessibility (Turner AW et al, 2022). (**d**) Location of rs604723 in critical nucleotides (in grey outlined boxes) of consensus transcription factor binding sequences for SRF and STAT6 as identified using the JASPAR 2022 database. (**e**) *IL5* association driven by lead eQTL rs7719499, exhibiting differences in expression by genotype but not sex or majority continental ancestry. (**f**) Regional association plot depicting variants in PAINTOR credible sets specific to BP (violet) or CAD (golden) GWAS annotations, mixFine only (light blue), mixFine and Paintor CS (green), and variants not in any credible set (light gray). (**g**) Variants of interest (rs7719499 and rs10065633) are indicated by blue lines and rs7719499 is highlighted in gray box, and corresponds to UCSC Genome Browser tracks indicating cell-type-specific chromatin accessibility (Turner AW et al, 2022). (**h**) Location of rs7719499 in critical nucleotides (in grey outlined boxes) of consensus transcription factor binding sequences for RUNX3 and nearby IRF4 (+10bp), as identified using the JASPAR 2022 database.

*IL5* encodes for Interleukin 5, an inflammatory cytokine with no eQTLs reported in any GTEx tissue but highly significant eQTLs in both STARNET arterial tissues (**Figure S6a-c**). Lead eQTL rs7719499 lies 75kb downstream of *IL5* adjacent to a plasma-specific chromatin accessible region for human coronary artery, proximal to *IRF1* (lead eQTL rs72797327, *p*_BH_=1, **Figure 4e-g**). Rs7719499 is also predicted to alter a RUNX3 transcription factor binding motif and is proximal to an IRF4 motif, suggesting putative cis-regulatory mechanisms in plasma cell types (**Figure 4h**). While *IL5* is expressed at low levels across most tissue types and coronary cell types (**Figure S6d**), it is significantly upregulated in aortic tissues from patients with CAD compared to controls (**Figure S6e, Table S16**).^44^

### Sensitivity analysis in European-ancestry subpopulation

We also assessed whether colocalization or fine-mapping could be improved by restricting our sample to European-ancestry individuals (n=87), who are vastly overrepresented in published genetics and genomics phenotypic studies and methods development. We identified 1,311 eGenes (16% discovery eGenes; 79% eGenes in the overall analysis, **Table S17**). With regard to generalization of published arterial eQTLs, 983 eGenes (75%) also had eQTLs in a GTEx or STARNET arterial tissue (46 of which were not significant eGenes in the overall mixQTL analysis, **Table S18**). In terms of colocalization, 51 eGenes colocalized to at least one GWAS signal. This represents only 47% compared to the overall sample, and fewer than would be expected in a similarly sized subset of a homogeneous study group (**Table S19**). Given the similarity in genetic architecture between this subset of our study population and GTEx and published GWAS, the reduction in associations across all analyses reinforces the benefits of including as many participants as possible to maximize statistical power.

### Coronary artery splicing QTL discovery

Differential isoform expression affects a wide array of complex diseases, and genetic variants affecting splicing events have been shown to be a major and distinct source of regulation underlying disease phenotypes.^45^ However, tissue-specific transcript specificity and isoform switching are not detected through eQTL methods unless total expression is affected. To identify genetic contributions to isoform-specific expression, we therefore evaluated genetic associations with 132,373 splice junctions in 14,815 genes. We identified 3,590 sQTLs (*p*_BH_<0.05) in 1,690 sGenes (**Figure 5a**, **Table S20**). Only 16% of sGenes were also identified as eGenes using either method (**Figure 2b**), pointing to the importance of evaluating isoform-specific in addition to total gene expression. Lead sQTLs for 61 splice junctions were annotated as splice region variants or splice acceptors in SnpEff, and nine discovery sGenes had protein-coding variants as lead QTLs (**Figure 5b**).

**Fig 5.**
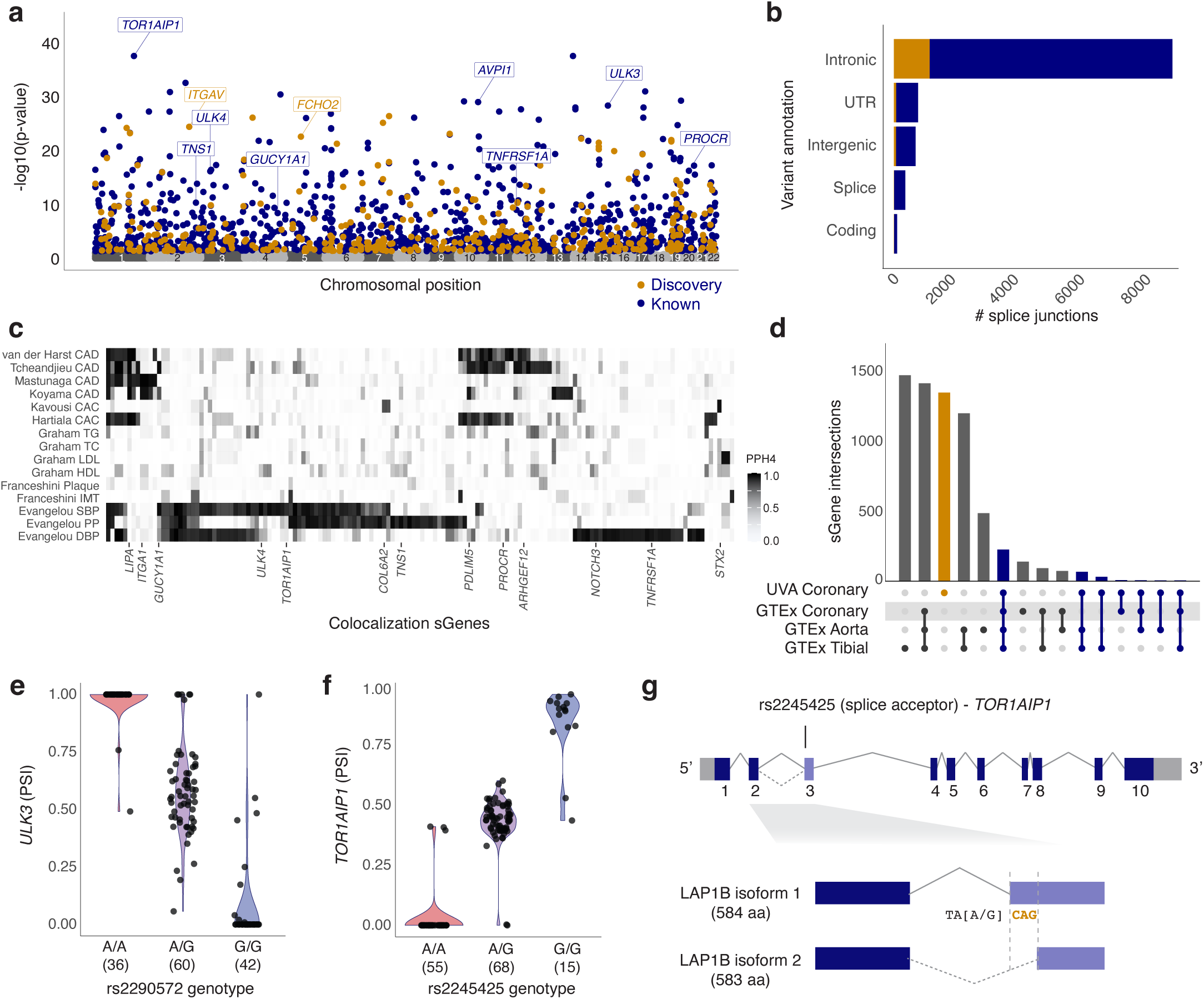
Coronary artery sQTL overview and characterization. (**a**) Manhattan plot of lead sQTLs. Navy blue and orange dots represent reported and discovery sGenes (pBH<0.05), respectively; gray dots represent non-significant genes. (**b**) SNPEff sQTL-sGene annotations for lead sQTLs; navy blue and orange represent numbers of reported and discovery sGenes. (**c**) GWAS colocalization to sQTL associations: each row represents the coloc PPH4 of the study (first author last name) and relevant GWAS trait, with intensity of shading corresponding to a higher posterior probability of a shared association at that locus. Each column represents one protein-coding gene with a PPH4;;0.8 in at least one GWAS. (**d**) Upset plot of generalization of GTEx arterial sQTLs. Black bars represent GTEx-specific sGenes; orange bars represent generalized sGenes, and the blue bar represents previously unreported sGenes. (**e,f**) Percent spliced in of *ULK3* and *TOR1AIP1* exons by genotype of their lead sQTLs (rs2290572 and rs2245425, respectively) in our study (left) and GTEx (right). (**g**) Schematic of *TOR1AIP1* gene and alternatively spliced isoforms, showing location and effect estimates for top splice acceptor variant (rs2245425) identified in our samples. rs2245425-G creates TAGCAG splice acceptor sequence at 3’ end of intron-exon 3 junction. Spliced CAG nucleotides (orange) encoding alanine amino acid distinguishes LAP1B isoform 1 and 2.

Colocalization of sQTL data exhibited a similar pattern to eQTLs, with a small subset of overlapping colocalized signals shared between CAD and BP trait GWAS and a higher number of quantitative trait associations (**Figure 5c**). No lead sQTLs overlapped directly with regions of open chromatin for any cell types. Compared to published arterial sQTL data, 871 (52%) were also sGenes in GTEx coronary artery, while 875 sGenes had no reported sQTLs in GTEx AOR, COR, or TIB tissues (**Figure 5d, Table S20**). There was overlap between genetically regulated gene expression and splicing: 297 eGenes were also sGenes, 23 of which shared the same lead variant (**Table S21**). Despite a similar pattern of colocalization, none of the eQTLs for shared associations colocalized with tested GWAS traits, affirming the likely different causal mechanisms underlying genetic effects on gene expression compared to splicing activity.

### Splicing contributes to CAD-relevant gene regulation

We present two plausible candidates for functional splicing effects in the coronary artery: *ULK3* and *TOR1AIP1*. *ULK3* is a broadly expressed serine/threonine kinase exhibiting autophosphorylation activity.^46,47^ The *ULK3* locus has also been associated with various CAD-relevant traits, including blood pressure, total cholesterol, and estimated glomerular filtration rate.^18,48–50^ Of thirteen junctions tested in our study, common missense variant rs2290572 was the lead sQTL for both significant associations: chr15:74837435:74837757 and chr15:74837435:74837751 (**Figure 5e**). Rs2290572 was also significantly associated with chr15:74837435:74837751 in GTEx tissues with directional consistency (**Figure S7a**). The lead sQTL for GTEx arterial tissues, rs12898397, is in high LD with rs2290572 in reference populations (**Table S11**), and represents a two-codon difference in an MIT domain in the fourteenth exon for which the T allele is predicted by SpliceAI to cause loss of a splice donor (Δ score=0.67, https://spliceailookup.broadinstitute.org).51 Rs12898397 is our lead eQTL (*p*_BH_=1.3E-14) and its contribution to isoform specificity makes it a clear candidate for functional follow-up.

TOR1AIP1 (Torsin 1A-interacting protein 1) is a broadly expressed lamin-binding protein which localizes to the inner nuclear membrane. Causal *TOR1AIP1* variants have been found for several autosomal recessive disorders, including limb-girdle muscular dystrophy with cardiac failure.^52,53^^52,52^ Located in the first intron, splice-acceptor variant rs2245425 is the lead sQTL for both significant splice excision events (of six tested in our study; chr1:179884769:179889313 and chr1:179884769:179889310), exhibiting nearly binary effects on the PSI of the third exon in coronary artery (**Figure 5f,g**). Rs2245425 is also the lead sQTL of GTEx arterial tissues (**Figure S7b**); *TOR1AIP1* has no eQTLs in our study or any GTEx artery tissue, pointing to splicing rather than transcription levels as the main effect of genetic regulation for this gene.

## Discussion

We report the first coronary artery eQTL mapping study accounting for local ancestry and allele-specific expression. Our study sample’s representation of both ancestral diversity and phenotypic heterogeneity allowed us to capture eQTLs that likely affect vessel wall integrity and maintenance throughout the lifecourse. The disruption of these transcriptional regulatory networks may be critical to plaque progression in coronary artery atherosclerosis. Our approach maximizes the likelihood of identifying eGenes in a rarely available tissue type, providing a roadmap for future QTL studies of diverse populations.

While tissues from patients with advanced CAD are useful for therapeutic development, profiling gene expression changes in subclinical CAD patients with multiple risk factors may point to avenues to prevent lesion progression. For example, molecular pathways for the strong epidemiologic association between blood pressure and CAD^54^ are incompletely described, while our work importantly begins to elucidate these mechanisms by highlighting a subset of eGenes colocalizing to GWAS findings of both traits. Furthermore, eGenes colocalizing to one GWAS trait can be prioritized for functional characterization, particularly those exhibiting cell-type specificity– e.g., Heparanase 2 (*HPSE2*), is enriched in SMCs in coronary artery and for which our eQTL colocalizes with SBP and DBP.^30^ *HPSE2* would not be a strong candidate gene in a QTL study focused on disease outcomes rather than intermediate phenotypes, but recent characterization of this extracellular matrix protein in endothelial maintenance suggests a contribution to vascular function during inflammation.^55,56^

Statistical fine-mapping analyses may prioritize causal genes and mechanisms of CAD loci, and we highlight example eGenes *TBX20* and *TCF21*. Here we identify the first genetic evidence of *TBX20* regulation in the human coronary artery, filling in a key knowledge gap in characterizing this gene in atherosclerosis. *TBX20* is required for normal cardiovascular development and was recently identified as regulating *PROK2*, a critical component of angiogenesis.^57^ *TBX20* has been implicated in causal pathways for congenital heart defects and continuous traits involving the great vessels.^58–60^ However, the role of *TBX20* in atherosclerosis has been minimally investigated, despite expression in arterial tissues and a well replicated CAD GWAS signal.^5,10,11^ Conflicting evidence about cell-type-specific expression of *TBX20* related to vascular function and neointimal hyperplasia^61^ demand functional characterization of regulatory elements in this locus that may modulate vessel wall pathways both during development and in a diseased state. Our *TCF21*/*TARID* findings also build on existing work by our lab and others characterizing an established CAD locus.^13,62–64^ This relationship is supported by research demonstrating regulation of *TCF21* expression by long non-coding RNA *TARID* in the context of CAD via promoter demethylation.^65,66^ The mural cell-enriched expression and chromatin accessibility of both genes, as well as overlapping association signals, compel a deeper look into the potential co-regulation of *TCF21* and *TARID* during SMC phenotypic transition.

It is now well appreciated that a large fraction of candidate regulatory variants are predicted to function via transcription factor binding-independent mechanisms, requiring comprehensive fine-mapping of candidate loci to prioritize likely causal variants.^20^ Our results expand the current focus on disease outcomes such as CAD by incorporating intermediate phenotype summary statistics into fine-mapping analyses. Using this approach, we narrowed colocalized GWAS signals to credible sets of candidate variants, highlighting *ARHGAP42* and *IL5*. The *ARHGAP42* locus is associated with blood pressure and cIMT,^67^ and gene expression changes affect smooth muscle cell contractility.^68^ Multi-omic fine-mapping revealed two candidate causal SNPs, of which the rs604723 risk allele has been shown to generate a cryptic SRF binding site to increase expression.^42^ We also highlight a putative STAT6 binding site created by the risk allele, suggesting a potential IL-4/IL-13-mediated activation of SMCs, both of which are normally lowly expressed in the coronary artery.

Another candidate gene, *IL5*, identified through our eQTL discovery and fine-mapping analyses, resides in a gene-dense SBP GWAS locus.^69^ While IL5 is reported to function in Th2 cells or eosinophils associated with atherosclerosis progression,^70–72^ its precise role and regulation in the coronary artery remains unknown. Notably, we did not identify eQTLs for neighboring inflammatory genes *IL4*, *IL13*, and *CSF2*,^73^ despite their potentially shared functions and regulatory mechanisms.^74,75^ Aorta-specific upregulation of *IL5* in STARNET CAD cases supports a potential disease-specific effect for *IL5* in multiple arterial tissues.^44^ Our lead eQTL, rs7719499, overlaps a plasma cell-specific chromatin accessibility peak in the coronary artery. Given the known influence of inflammatory cytokines on endothelial and SMC activation, future studies are warranted to investigate the immune cell derived IL5 mediated vascular wall injury in cell and animal models.

Finally, we demonstrated distinct genetic contributions for expression and splicing activity in the coronary artery. The relationship between isoform specificity and overall expression is complex.^76,77^ We observed a modest sharing of eGenes and sGenes (14%) and fewer than 10% overlap with regard to GWAS colocalization, supporting orthogonal effects of genetic variation on transcription compared to isoform regulation. We explored two sGenes with potential roles in coronary artery disease–*ULK3* and *TOR1AIP1*–both of which exhibit tissue-specific isoforms and strong evidence of genetically regulated splicing. Causal *TOR1AIP1* mutations have been reported for monogenic dystrophic developmental disorders via dysregulation of necessary protein complex formation at the inner nuclear membrane,^52,78^ suggesting a cardiac-specific function for one or more transcripts. The *TOR1AIP1* sQTL regulates the inclusion of three base pairs, adding an alanine residue to the third exon (position 185). While this variant has not been associated with a specific disease, its high allele frequency suggests differential isoform expression may have an important functional role, particularly given the lack of *TOR1AIP1* eQTLs. Regarding *ULK3*, though mainly described in the context of cancers,^79^ this gene may mediate vascular disease through dysregulation of autophagy and interactions in the Shh signaling pathway.^80,81^ Single-cell sequencing data reveal a strong enrichment for *ULK3* expression in mast cells, for which conflicting evidence exists regarding effects on CAD-related outcomes via histamine production.^82,83^ Although the *ULK3* locus has also been reported in BP and cholesterol GWAS, the published associations are located downstream of the *ULK3* coding region and are statistically independent of our sQTL and eQTL.^9,48,69^ Our findings for both transcriptional and isoform-specific regulation of this promising candidate gene by variants with no GWAS signal point to the importance of considering multiple ‘omics datasets both together and independently when evaluating highly complex diseases and traits.

Overall, these findings emphasize the importance of context in interpreting genetic associations with disease. Characterizing loci within a comprehensive genomic and physiologic setting helps prioritize top candidate genes with unique genomic profiles relevant to atherosclerosis disease processes. While coronary artery tissue remains the most relevant single tissue type for prioritizing CAD candidate genes, the gene expression program changes during atherosclerosis likely involve complex, multi-tissue, and multi-cellular gene regulatory networks. Multi-tissue network analyses may further resolve underlying paracrine signaling pathways and regulatory mechanisms for eGenes without an obvious role in predominant intimal or medial cell types.^84^ For instance, inflammatory processes driven by cytokine signaling may be difficult to detect in target cell types, but significantly different expression of *IL5* specifically in the aorta in STARNET cases compared to controls provides both validation of our approach and options for identifying other genes functioning in the same molecular pathway.^44^

It is worth noting limitations both common to eQTL studies and unique to our approach. First, restricting to 5% MAF variants limited detection of ancestry-specific associations; previous work has shown that effectively capturing lower frequency variants increases discovery both across and within ancestral populations.^85^ A second limitation relates to interpretability of our colocalization and fine-mapping results based on genetically homogeneous public datasets.^17^ Individuals with genetic ancestry from Africa and the Americans in particular continue to be severely underrepresented in both GWAS and ‘omics reference datasets. Genetic homogeneity in study populations not only prevents identification of ancestry- or haplotype-specific associations, but also limits generalizability of global associations when fine-mapping is restricted to genetic architecture from a single ancestry group.^86,87^

In contrast, a major strength of our work is demonstrating the feasibility and promise of incorporating local genetic ancestry and allele-specific expression into eQTL analyses to discover disease-relevant genes/pathways. We acknowledge the caveats of using continental genetic reference populations, which lack granularity and global representation. However, our study design increased statistical power while reducing the unnecessary exclusion of individuals based on ancestry or ethnicity, providing a guideline for implementation in future studies. Further, incorporation of single-cell chromatin accessibility datasets in coronary artery complemented our eQTL-based gene prioritization approach to nominate cis-regulatory mechanisms underlying complex diseases associations.^37^ Finally, our findings benefited from comparing epigenomic and genetic annotations, providing a more tenable suite of candidate variants for future functional work. Our multiple phenotype fine-mapping approach will be particularly relevant as the field moves toward functional characterization of disease-associated lncRNAs and splice isoforms, for which traditional metrics such as evolutionary conservation cannot be consistently applied.

In summary, we present the first genetically diverse evaluation of coronary artery gene expression across a phenotypic spectrum of atherosclerosis. Our inclusive study design with respect to ancestry and robust pipeline facilitated the discovery of atherosclerosis-associated genes with plausible functional roles in the vascular wall. Molecular characterization of these genes in environments representing subclinical atherosclerosis will improve the identification of therapeutic targets for CAD patients.

## Methods

### Ethics Statement

All research described herein complies with ethical guidelines for human subjects research under approved Institutional Review Board (IRB) protocols at Stanford University (#4237 and #11925) and the University of Virginia (#20008), for the procurement and use of human tissues and information, respectively.

#### 1. Sample Acquisition

Freshly explanted hearts from heart transplant recipients were obtained at Stanford University under approved Institutional Review Board protocols and written informed consent. Hearts were arrested in cardioplegic solution and rapidly transported from the operating room to the lab on ice. The proximal 5-6 cm of three major coronary vessels (left anterior descending (LAD), left circumflex (LCX), and right coronary artery (RCA)) were dissected from the epicardium on ice, trimmed of surrounding adipose and adventitia, rinsed in cold phosphate buffered saline, and rapidly snap frozen in liquid nitrogen. Similarly, aortic root and left ventricular free wall tissues were also processed and stored at -80C until processing. Throughout the manuscript, these explanted tissues will be referred to as “Explants”. Normal coronary artery, aorta, and left ventricle tissues were obtained by Stanford University (from Donor Network West and California Transplant Donor Network) from non-diseased donor hearts rejected for orthotopic heart transplantation, procured for research studies and were treated following the same protocol as the explanted hearts. The collected tissues will be referred hereby as “Donors”. Tissues were de-identified and clinical information (e.g., ICD-10 codes for primary and secondary diagnoses) was used to classify normal, ischemic and non-ischemic hearts. Frozen tissues were transferred to the University of Virginia through a material transfer agreement and Institutional Review Board approved protocols.

#### 2. DNA Genotyping

##### 2.1 Genomic DNA Isolation

Approximately 20-25 mg of frozen left ventricle or coronary artery tissue was used to isolate genomic DNA for each donor sample following the manufacturer’s protocol (Qiagen DNeasy Blood and Tissue Kit, cat# 69504).

##### 2.2 Genomic DNA sequencing

Genomic DNA samples for all donors in the study were diluted using TE buffer to [5-15 ng/µL] in skirted 96-well PCR plates. Plates were sealed and shipped to Gencove (New York, USA) for 0.4X low-pass genomic DNA sequencing.

##### 2.3 Genomic DNA Sequence Processing

###### Phasing and imputation

Unphased low-pass whole genome sequencing files were provided by Gencove in build b37. VCFs provided by Gencove included just over 38 million variants imputed to 1000G using a proprietary pipeline–imputation quality scores were not provided. Samples were phased and re-imputed to 1000G phase 3 b37 reference panel using Beagle with impute=true and gp=true options–phasing was necessary for downstream analyses but no additional variants were imputed.^88,89^

###### Liftover of Genomic Coordinates

Phased autosomal VCFs were lifted over from b37 to hg19 to hg38 using Picard (“Picard Toolkit”, 2019, Broad Institute. GitHub Repository: http://broadinstitute.github.io/picard/). After excluding approximately 10,000 variants that could not be mapped, approximately 38 million total variants were available for consideration in analyses.

##### 2.4 Principal component estimation

We calculated ancestral principal components using the R package SNPrelate.^90^ Briefly, SNPRelate uses LD pruning to restrict genotype data to ∼500,000 independent biallelic SNPs with a MAF >1% across the autosomal chromosomes. We used all participants from 1000 Genomes Phase 3 as a reference panel given the diversity of our sample.^91^ Eigenvectors (EVs) 1 through 3 demonstrated clustering and correlation with Gencove-assigned majority continental ancestry, but subsequent EVs were driven by one or a small number of individuals (**Figure S1**). Analyses adjusting for global ancestry were therefore restricted to the first three EVs.

##### 2.5 Local ancestry estimation

Broad ancestral diversity, along with admixture implied by Gencove-reported regional ancestry estimates, suggested that adjustment for local ancestry (LA) might improve discovery compared to global EVs. We adapted the local ancestry pipeline developed by Alicia Martin (https://github.com/armartin/ancestry_pipeline), including incorporation of several of Dr. Martin’s Python scripts.^92^ We used RFmix2 (https://github.com/slowkoni/rfmix) to calculate ancestry from one of five continental reference populations^93^; ancestry was recorded as “missing” for a particular region if >90% probability of concordance with one reference population could not be attained. For computational efficiency, we randomly selected genotypes representing continental ancestries from each superpopulation for 1,200 total 1000 Genomes participants: 400 from AFR (African), and 200 each from AMR (Indigenous to the Americas), EAS (East Asian), EUR (European), and SAS (South Asian) superpopulations.^91^ Because the most recent complete reference dataset with genetic map data for 1000G is b37 (corresponding to build hg19), we performed LA estimation and all downstream analyses using hg19 positions and gencode ‘v37lift37’ annotations (see “Local Ancestry Adjustment”).^94^

#### 3. Bulk RNA-seq

##### 3.1 Coronary artery tissue processing and RNA Isolation

Coronary artery samples were selected for RNA sequencing based on several criteria, including tissue availability (>50mg) and disease status (prioritizing capturing a range of phenotypes). Total RNA was extracted from frozen coronary artery segments using the QIAGEN miRNeasy Mini RNA Extraction kit (catalog #217004). As could be expected, ease of sample processing under liquid nitrogen varied along with differing levels of calcification by sample. Approximately 20 mg of frozen tissue pulverized using a pre-chilled mortar and pestle under liquid nitrogen was added to 1.5 mL RINO tubes (Next Advance, SKU TUBE1R5-S) which were stored on dry ice. Tissue powder was then further homogenized in Qiazol lysis buffer using stainless steel beads in a Bullet Blender (Next Advance) homogenizer, followed by column-based purification according to the manufacturer’s instructions. RNA concentration was determined using Qubit 3.0 and RNA quality was determined using Agilent 4200 TapeStation. Samples from three to five donors were processed per day by one of three individuals. Samples with RNA Integrity Number (RIN) >4.5 and Illumina DV_200_ values >75 were included for library construction.

##### 3.2 RNA Library Sequencing

Total RNA libraries were constructed using the Illumina TruSeq Stranded Total RNA Gold kit (catalog #20020599) and barcodes were added to RNA libraries using the IDT for Illumina - TruSeq RNA Unique Dual (UD) Indexes (96 Indexes, 96 Samples) Kit (IDT, Illumina, catalog #20022371. This library preparation captured coding RNAs and some noncoding RNAs, while depleting ribosomal RNAs. After re-evaluating library quality using TapeStation, individually barcoded libraries were sent to Novogene for next generation sequencing. After passing additional QC, libraries were multiplexed and subjected to paired end 150 bp read sequencing on an Illumina NovaSeq S4 Flowcell to a median depth of 100 million total reads (>30 G) per library.

##### 3.3 RNA-seq Mapping

Our RNA sequencing QC pipeline and scripts pertaining to all analyses described can be publicly accessed at: (https://github.com/MillerLab-CPHG/CAD_QTL)

###### RNA-seq Read Mapping and Quality Control

The raw passed filter sequencing reads obtained from Novogene were demultiplexed using the bcl2fastq script. Read quality was assessed using FastQC (https://www.bioinformatics.babraham.ac.uk/projects/fastqc/, version 0.11.9) and the adapter sequences were trimmed using Trim Galore version 0.6.5 (https://www.bioinformatics.babraham.ac.uk/projects/trim_galore). Reads with low average Phred scores (<20) were also removed, after which all samples passed the quality control analysis were considered for inclusion. Trimmed reads were mapped to the hg38 human reference genome using STAR v2.7.3a according to the GATK Best Practices for RNA-seq.^95^ To increase mapping efficiency and sensitivity, novel splice junctions discovered in a first alignment pass with high stringency, were used as annotation in a second pass to permit lower stringency alignment and therefore increase sensitivity. PCR duplicates were marked using Picard and WASP was used to filter reads prone to mapping bias. Total read counts and RPKM were calculated with RNA-SeQC v1.1.8 using default parameters and additional flags “-n 1000 -noDoC -strictMode” and GENCODE v32 reference annotation. Transcript and isoform expression levels were estimated using RSEM (v1.3.3).^96^

###### Detection of RNA-seq Sample Swaps

Using the previously known genotypes for an individual from our cohort, we used verifyBamID ^97^ to check the possibility of whether the reads were contaminated or sample swaps occurred, we identified four swapped samples. To cross check these swaps, we ran NGSCheckMate, which uses a depth-dependent correlation model of allele fractions of known single -nucleotide polymorphisms (SNPs) to identify samples from the same individual.^98^ Consistent with VerifyBamID, we found four swapped samples, two of which we rematched and two of which were duplicates and therefore excluded from downstream analysis.

###### RNA sequencing data phasing for allele-specific expression

In order to capture allele-specific expression within RNAseq data, we phased all reads using RASQUAL.^99^ Haplotype phasing of RNA sequencing reads was performed using phASER v1.1.1.^100^ Known sites for allelic mapping bias and HLA genes were excluded because of their high mapping error rates and introduction of bias using the ENCODE Unified GRCh38 Blacklist (as of 5-5-2020).^101^ The phASER pipeline was performed using the guided tutorial with additional flags; “--paired_end 1 --mapq 255 --baseq 10”. Generation of the haplotype expression quantifications was performed with the companion tool called “phASER Gene AE” using the standard pipeline and the GENCODE v36 GRCh38 gene coordinates for haplotypic expression calculation.^94^

###### Multidimensional Scaling

We performed nonparametric clustering in transcriptomic space to determine whether the transcriptomes of our coronary artery samples were unique from GTEx tissues as well as previously published HCASMC transcriptomes. We performed multidimensional scaling on log-transformed RPKM values (excluding genes in our study which did not have ≥0.1 RPKM in ≥10 individuals) of all GTEx tissues, our samples, and HCASMC cells as previously described using Kruskal’s non-parametric method. The 54 unique GTEx tissue sites were grouped into 27 broad tissue categories as previously described^13^.

#### 4. Quantitative Trait Loci Identification and Characterization

##### 4.1 eQTL Analysis

###### PEER Factor calculation and optimization

Probabilistic Estimation of Expression Residuals (PEER) was used to account for unmeasured confounders related to RNA sequencing (e.g., batch effects).^102^ No covariates were included. Based on recommendations from the original publication regarding sample size and statistical models, we expected to incorporate up to five PEER factors in all models. In order to determine how the number of PEER factors may affect our results, we selected a random subset of 400 genes on chr17 (∼30% of annotated lncRNAs or protein-coding genes on that chromosome). We then performed linear regression using QTLtools (described below but with an allele frequency cutoff of 1% rather than 5%) with no additional covariates and compared the number of eGenes with an FDR <0.05 adjusting for 1, 2, 3, 5, 8, 10, 15, 20, and 25 PEER factors (**Figure S1**). The number of eGenes was not meaningfully affected by the number of PEER factors, hence we continued with five as expected.

###### Regression analyses

To maximize true positive associations and minimize Type I error, we excluded variants that did not meet the following criteria: biallelic single-nucleotide variants with a minor allele frequency >5% and HWE p-value >1E-6 (http://samtools.github.io/bcftools/bcftools.html). Approximately 6,100,000 variants were available for inclusion in each analysis. Bed files were generated using the criteria implemented by GTEx: all lncRNA and protein-coding genes exceeding 0.1 TPM for at least 20% of samples were included^29^. Genes were annotated with name, genomic coordinates, and strand using Gencode v32 (https://www.gencodegenes.org/human/release_32.html) for hg38 analyses and gencode v37liftb37 (v37 lifted “under” from hg38 to hg19 positions, https://www.gencodegenes.org/human/release_38lift37.html) for hg19 analyses.^94^ Due to strong selection and high likelihood of population stratification contributing to false-positive identification of genes in the MHC region which is under high selective pressure, *HLA* genes and other MHC components were not considered candidates for fine-mapping in downstream analyses.

###### Statistical significance reporting

For all regression analyses, the following standards will be used for reporting p-values throughout the manuscript. For single variant association tests, p_nom_ refers to the p-value reported in the “pval_meta” column for mixQTL or the 12^th^ output column for QTLtools run under the nominal pass model. For variants that were not lead QTLs, *p*_adj_ refers to the Bonferroni-corrected p_nom_, where the correction is adjusting for the number of SNPs tested for each gene or splice junction respectively. For lead QTLs, *p*_BH_ refers to the value obtained by correcting for the total number of genes or splice junctions tested within each method using the Benjamini-Hochberg FDR correction applied to the *p*_adj_ for mixQTL analyses and the 19^th^ output column for QTLtools analyses.

###### Local ancestry adjustment

Local-ancestry-adjusted cis-eQTL analyses were performed using a new pipeline which incorporates QTLtools (a computationally efficient implementation of MatrixQTL which allows adjustment for SNP-level covariates).^103,104^ Models adjusted for age, sex, local ancestry, and five PEER factors. Because local ancestry designations occur at the SNP level and cannot be included as traditional covariates in genome-wide eQTL regression analyses, proportions of estimated continental ancestry for each individual were calculated for each gene, and gene-specific covariate and bed files were used in QTLtools. Local ancestry interpolation scripts as well as scripts to run each gene individually and select lead SNPs with adjusted p-values can be found on the lab GitHub repository referenced above.

Permutation pass mode was run in QTLtools using up to 100,000 iterations per gene based on the method described by Gay, et al,^25^ to generate a lead variant for each gene tested. “Nominal-pass” results were run for significant eGenes only to obtain a non-permuted p-value for each variant within 500kb up- or downstream of the transcription start site (TSS). Isoforms were not evaluated separately, therefore the TSS used for each gene was the most upstream TSS for transcripts with multiple annotated isoforms.

###### Combined global-ancestry-adjusted and allele-specific expression analysis

We implemented the R package ‘mixQTL’ (https://github.com/hakyimlab/mixqtl) to identify eQTLs incorporating allele-specific expression in our data.^24^ All mixQTL analyses were performed using genome build hg38; RNA transcripts were mapped to Gencode v32 (https://www.gencodegenes.org/human/release_32.html). Briefly, mixQTL tests for an association between a genotype and total read count; an association between specific alleles and corresponding haplotype expression (allele-specific expression); and the meta-analysis of both scores when inclusion criteria are met for both methods. MixQTL inputs include phased genotypes, total read counts, allele-specific read counts, and covariate information. Because mixQTL has to be run separately for each gene and does not have a way to account for missing data, all data frames (covariate, haplotype, and expression) would need to be re-generated for each gene in the course of running the R script to account for “missing” values for any covariate, haplotype, or expression value for one or more individuals. We did not have missing data for any included covariates or genetic variants. Most genes with any missing expression values were lowly expressed, therefore we do not expect inflation that would not be accounted for by adjusting the resulting p-values, and set all missing values in the phased RNAseq data to zero. We adjusted the example script on the mixQTL Github (https://github.com/liangyy/mixqtl) to limit our window to variants within 500kb of the transcription start sites of each gene for a maximum window size of 1Mb, and analyzed associations all genes with ≥0.1 mean TPM. MixQTL models included three ancestry EVs, age, sex, and five PEER factors as covariates.

###### European-ancestry-specific mixQTL sensitivity analyses

Most statistical fine-mapping methods and publicly available genomics references continue to over-represent ancestrally homogeneous European-ancestry populations. To evaluate whether discovery or replication could be improved despite a meaningful decrease in sample size, we performed mixQTL and downstream analyses in a subset of our study population restricted to individuals with 100% European ancestry (n=79) with follow-up methods as described above.

##### 4.2 Splicing analysis

###### Generation and processing of BAM and junction files for splice QTL analysis

We aligned to the human genome the FASTQ files from RNAseq data of 138 ancestrally diverse heart transplant patients using the aligner tool STAR (version 2.7.2b) in 2-pass mode.^105^ A STAR index was generated using Gencode v37 annotations and the UCSC hg38 reference genome. BAM files were generated using the --twopassmode basic and --outSAMtype BAM SortedByCoordinate options with a minimum overhang of 8 bp for spliced alignments. Additionally, the WASP correction option, --waspOutputMode SAMtag, was used to mitigate allelic mapping bias.^106^ The generated BAM files were further indexed using the option --index from the package Samtools.^107^ Next, junction files were generated using the package RegTools.^108^ The option “-junctions extract” was used with the following parameters: (1) minimum intron size of 50 bp, (2) maximum intron length (100 kbp), (3) minimum overlap between junctions of 8 bp, and (4) leaving the strand specificity as ‘unstranded’ to analyze both positive and negative strands. We removed unknown or unwanted chromosomes from junction files.

###### LeafCutter

QTL input files were generated using the modified LeafCutter-GTEX pipeline.^109,110^ Junction files were used as input to generate a count matrix of intron excision ratios. Clusters of alternatively spliced introns were identified using split reads that mapped with a minimum of 8 bp into each exon. Singleton introns that did not cluster with other introns were discarded. LeafCutter iteratively analyzed each intron cluster and removed introns in two ways: (1) introns with fewer than 50 reads across all samples, or (2) intron reads present in less than 0.1% of the total number of reads in the entire cluster. LeafCutter re-clustered introns and only included those with a maximum length of 100,000 bp. The count matrix was further processed to (1) generate a BED file and its index in a format compatible for QTL mappers, such as QTLtools; and (2) to calculate splice principal components. The LeafCutter protocol specifically outputs percent spliced in (PSI) for each splice junction, which adjusts for splicing events with overlapping start or stop positions to account for gene-level variation.

###### sQTL association

Genetic variants affecting the quantity of proximal splice junctions (cis-sQTLs) were estimated using QTLtools as described above, with age, sex, and four ancestry principal components included in the model. Nominal-pass was used to obtain non-permuted p-values for each variant within 250kb up- or downstream of the LeafCutter-generated BED file of splice junction expression. 100,000 permutations were run with a permutation pass to generate an adjusted p-value for each lead sQTL.

##### 4.2. Characterization of coronary artery QTLs

###### Generalization of previously published eQTLs in our study

We compared LA and mixQTL eGenes to published eQTL datasets in relevant tissues. Specifically, we assessed whether eGenes in our study also had significant eQTLs–and whether our lead variants were correlated using European-ancestry LD–in GTEx v8 coronary, thoracic aorta, and tibial artery tissues;^29^ and STARNET internal mammary artery and aortic root tissues.^44^ Due to differences in annotation, overlap between each dataset and genes meeting our inclusion criteria was identified using the maximum set of gene names or primary ENSG identifiers (excluding post-decimal identifying numbers).

###### Annotation of eQTLs and sQTLs

To identify potential causal mechanisms of QTLs, we annotated lead eQTLs and sQTLs to their respective eGenes and sGenes using the hg38kg database in snpEff.^111^ SnpEff reports variant annotations by gene (i.e., specific variant-gene annotations are not provided in the main output). For this reason, we utilized the verbose (-v) option and the log file to identify gene-variant-specific annotations to ensure the eQTL-eGene association was maintained. Corresponding scripts are available on our Github. We further evaluated annotations using the annotate_variation.pl script with Annovar, also for build hg38.^112^

###### Co-localization of eQTLs and sQTLs with GWAS Variants

We tested for colocalization of eQTLs with significant (p<5×10^-8^) published genetic associations in GWAS of coronary artery disease and myocardial infarction.^5,8–11^ Due to the phenotypic heterogeneity within our study population, we additionally evaluated quantitative traits that are traditional risk factors for CAD including blood pressure traits (systolic [SBP], diastolic [DBP], and pulse [PP] pressures),^69^ cholesterol traits (total cholesterol [TC], high-density lipoprotein [HDL], low-density lipoprotein [LDL], and triglycerides (reported as a log concentration, logTG])^18^, carotid artery calcification (CAC),^113^ and intima media thickness (IMT).^114^ The GWAS catalog (https://www.ebi.ac.uk/gwas) was most recently accessed on November 1, 2022. We considered PPH3 >0.8 to implicate two independent causal SNPs associated with eQTL and GWAS association signals, and PPH4 >0.8 to support evidence of a shared causal variant.^115,116^

To evaluate the contribution of QTL associations with causal pathways for CAD, we also used coloc to colocalize CAD/MI GWAS and quantitative trait summary statistics. To annotate likeliest causal genes for colocalized associations between two GWAS, we annotated variants with the highest posterior probability of causality in associations with PPH4>0.8 to GTEx arterial tissues (AOR, COR, or TIB).

###### Summary-data-based Mendelian Randomization (SMR)

SMR evaluates evidence for statistical pleiotropy by comparing summary statistics from GWAS (in our study: CAD/MI and blood pressure traits) to eQTL results from expression data in relevant tissues; i.e., to test whether the association between a genetic variant and CAD-related phenotypes is mediated by gene expression in our study population^34^. We used the 1000G EUR population (excluding Finnish samples due to genetic distinction of that population, which was not represented in our samples) as our reference population. We incorporated GWAS summary statistics from the same studies used for coloc to perform SMR with mixQTL results (https://yanglab.westlake.edu.cn/software/smr). We were not able to evaluate either Japanese GWAS due to the large number of variants with allele frequency differences >30% between the GWAS study populations and ours.

##### 4.3 Fine-mapping of eGene association signals

###### mixFine

To identify independent associations and prioritize credible sets for discovery coronary artery eGenes, we used the command line implementation of mixFine (https://github.com/hakyimlab/mixqtl).24 Mixfine runs SuSiE internally to accommodate the possibility of multiple causal variants in a dataset where many of the input variables (SNPs) are likely highly correlated.^116^ Input files utilized by mixFine are the same as those for mixQTL and were prepared as described above.

###### Fine-mapping with GWAS and variant annotations

To combine the effects of GWAS association signals, eQTLs identified in our coronary artery tissue samples, and tissue-relevant variant annotations, we fine-mapped mixQTL eGene associations using fast Paintor (v3.0, https://github.com/gkichaev/PAINTOR_V3.0).117 Due to FastPaintor input requirements and the ancestral representation in published GWAS of relevant traits, we restricted published datasets included in these analyses to European-only ancestry. MixQTL results were combined with overlapping SNPs from GWAS summary statistics for two groups of traits–CAD/MI and blood pressure traits.^5,9,10^ For each eGene (+/-500kb from TSS), VCFs and LD matrices were generated with Plink, using hg38 1000G EUR samples excluding FIN samples (which are minimally represented in our population or published GWAS).^91,118,119^ We included ENCODE binary SNP annotations for chromatin accessibility (CTCF, H3K27ac, and H3K4me3 in coronary artery tissue from one 53yo female).^15^ To apply enriched enhancer regulatory activity in coronary artery tissue, we also used the activity-by-contact model (ABC; https://github.com/broadinstitute/ABC-Enhancer-Gene-Prediction). ABC scores were obtained from previous H3K27ac HiChIP and ATAC data in HCASMC and coronary artery from our previous work are publicly available (see “Data and Code Availability).^13,120^

###### Overlap with cell-type-specific chromatin accessible regions

To evaluate whether lead QTLs were likely to directly affect gene expression or splicing via altering chromatin accessibility, we intersected cell-type-specific snATAC data from 41 individuals in our study population with the positions of all lead QTLs.^37^ We used the Bedtools intersect function for this with the -C flag to provide output for all tested SNP-region pairs.^121^

## Data Availability

All raw and processed bulk RNA-sequencing data will be made available on the Gene Expression Omnibus (GEO) database (accession number:). Low-pass whole-genome sequencing-based genotyping data are available on dbGaP (accession code phs002855.v1.p1). The full summary statistics for the mixQTL eQTL analyses, as well as the local ancestry eQTL and the sQTL analyses are available here: https://doi.org/10.5281/zenodo.7581778
The single-cell RNA-seq datasets from coronary and carotid artery were re-analyzed and integrated from the original datasets available through GEO (accession numbers: GSE131778 (Wirka, et al.), GSE155512 (Pan, et al.), GSE159677 (Alsaigh, et al.) and https://doi.org/10.5281/zenodo.6032099 (Hu, et al.). The raw and processed single-nucleus ATAC-seq datasets are available through GEO (GSE175621 and GSE188422). The reprocessed and analyzed human scRNA-seq datasets are also available on PlaqView (https://plaqview.com). GTEx gene expression and eQTL data were obtained from the v8 portal website (https://gtexportal.org). STARNET gene expression, eQTL, and clinical trait enrichment data were obtained from dbGaP (accession number: phs001203.v2.p1) and are also available at http://starnet.mssm.edu. The HCASMC ATAC-seq and H3K27ac HiChIP data used to calculate ABC scores are available through GEO (accession numbers: GSE113348 and GSE101498).
All custom scripts used to generate the results are available on GitHub (https://github.com/MillerLab-CPHG/CAD_QTL). Detailed parameters for published software tools are also included in the Methods.

## Abbreviations

**General**
FDR: False discovery rate
GWAS: Genome-wide association study (or studies)
HCASMC: Human coronary artery smooth muscle cells
LA: Local-ancestry adjusted QTL analysis
QTL: Quantitative trait locus
SMC: Smooth muscle cell
SNP: Single nucleotide polymorphism

**Diseases and traits**
BP: Blood pressure
CAD: Coronary artery disease
DBP: Diastolic BP
HDL: High-density lipoproteins
LDL: Low-density lipoproteins
logTG: Log_10_ of triglycerides
MI: Myocardial infarction
PP: Pulse pressure
SBP: Systolic BP
TC: Total cholesterol

**Study-specific**
GTEx: Genotype-Tissue Expression Project
STARNET: Stockholm-Tartu Atherosclerosis Reverse Network Engineering Task
AOR: Aorta
COR: Coronary artery
MAM: Mammary artery
TIB: Tibial artery
1000G: 1000 Genomes Project
AFR,AMR,EAS,EUR,SAS: 1000G superpopulation abbreviations for participating individuals from populations in Africa, the Americas, East Asia, Europe, and South Asia, respectively.

**Gene names**
*AKT3*: AKT Serine/Threonine Kinase 3
*ARHGAP42*: Rho GTPase Activating Protein 42 (protein AKA GRAF3)
*HPSE2*: Heparanase 2
*IL5*: Interleukin 5
*LIPG*: Lipase G, endothelial type
*TARID*: TCF21 Antisense RNA Inducing Promoter Demethylation
*TBX20*: T-Box 20
*TCF21*: Transcription factor 21
*TOR1AIP1*: Torsin 1A Interacting Protein 1
*ULK3*: Unc-51 Like Kinase 3
*YY1AP1*: YY1-associated protein 1

## Data and Code Availability

All raw and processed bulk RNA-sequencing data will be deposited on the Gene Expression Omnibus (GEO) database. Low-pass whole-genome sequencing-based genotyping data are available on dbGaP (accession code phs002855.v1.p1). The full summary statistics for the mixQTL eQTL analyses, as well as the local ancestry eQTL and the sQTL analyses are available here: https://doi.org/10.5281/zenodo.7581778

The single-cell RNA-seq datasets from coronary and carotid artery were re-analyzed and integrated from the original datasets available through GEO (accession numbers: GSE131778 (Wirka, et al.), GSE155512 (Pan, et al.), GSE159677 (Alsaigh, et al.) and https://doi.org/10.5281/zenodo.6032099 (Hu, et al.). The raw and processed single-nucleus ATAC-seq datasets are available through GEO (GSE175621 and GSE188422). The reprocessed and analyzed human scRNA-seq datasets are also available on PlaqView (https://plaqview.com). GTEx gene expression and eQTL data were obtained from the v8 portal website (https://gtexportal.org). STARNET gene expression, eQTL, and clinical trait enrichment data were obtained from dbGaP (accession number: phs001203.v2.p1) and are also available at http://starnet.mssm.edu. The HCASMC ATAC-seq and H3K27ac HiChIP data used to calculate ABC scores are available through GEO (accession numbers: GSE113348 and GSE101498). All custom scripts used to generate the results are available on GitHub (https://github.com/MillerLab-CPHG/CAD_QTL). Detailed parameters for published software tools are also included in the Methods.

## Acknowledgements

This work was supported by grants from: the National Institutes of Health (grant numbers R01HL148239 and R01HL164577 to C.L.M; T32HL007284 to C.J.H; and R01HL125863 to J.L.M.B; R01HL130423, R01HL135093 and R01HL148167 to J.C.K.), the American Heart Association (grant number 20POST35120545 to A.W.T.; AHA909150 to J.V.M.; A14SFRN20840000 to J.L.M.B.), the Swedish Research Council and Heart Lung Foundation (grant number 2018-02529 and 20170265 to J.L.M.B.), the Fondation Leducq (grant number ‘PlaqOmics’ 18CVD02 to C.L.M., J.L.M.B.) and the Single-Cell Data Insights award from the Chan Zuckerberg Initiative, LLC and Silicon Valley Community Foundation (to C.L.M.). This work was also supported by fellowship grants from the Bench to Bassinet Pediatric Cardiac Genomics Consortium (PCGC) & Cardiovascular Development Data Resource Center (CDDRC) (to C.J.H.), as well as the UVA MSTP training grant (NIH T32GM007267, to W.F.M.). The authors would like to thank Yipei Song and Wesley Craig for assistance with scripting and troubleshooting; Catherine Robertson for assistance with fine-mapping study design; Katia Sol-Church and Yongde Bao for assistance with library preparation and sequencing; Peter Chiu, Paul Chang, A.J. Pedroza, Tiffany Koyano, Euan Ashley, Tom Quertermous, and all of the transplant recipients, heart donors, family members, study coordinators, and transplant procurement team at Stanford for coronary artery tissue procurement.

## Author contributions

We used CRediT taxonomy to determine the authors contributions. C.J.H., A.W.T., J.L.M.B., and C.L.M conceptualized the study. C.J.H., A.W.T., M.D.K., N.G.L. curated the data. C.J.H., A.W.T., M.D.K., N.B.B., R.M., L.M., J.V.M., performed formal data analysis. C.J.H., A.W.T., N.J.L., J.C.K., J.L.M.B., and C.L.M. acquired funding. C.J.H., A.W.T., and E.F., performed the experiments. M.D.K., N.B.B., R.M., E.F., D.W., and S.O-G. contributed to methods development. G.A. and S.O-G. performed project administration. A.W.T., L.M., N.G.L, J.V.M., W.F.M., M.K., P.A.P., S.W.vdL., N.J.L., J.C.K., J.L.M.B., and C.L.M. contributed biospecimens, datasets, scripts or other resources. C.J.H., and W.F.M. contributed software tools. C.L.M supervised the project. L.M., J.V.M., and J.L.M.B. contributed to validation of the results. C.J.H., A.W.T., M.D.K., N.B.B., J.V.M., and C.L.M. contributed to data visualization. C.J.H., A.W.T., and C.L.M. prepared the manuscript draft. All authors reviewed and edited the manuscript.

---

**Fig S1.**
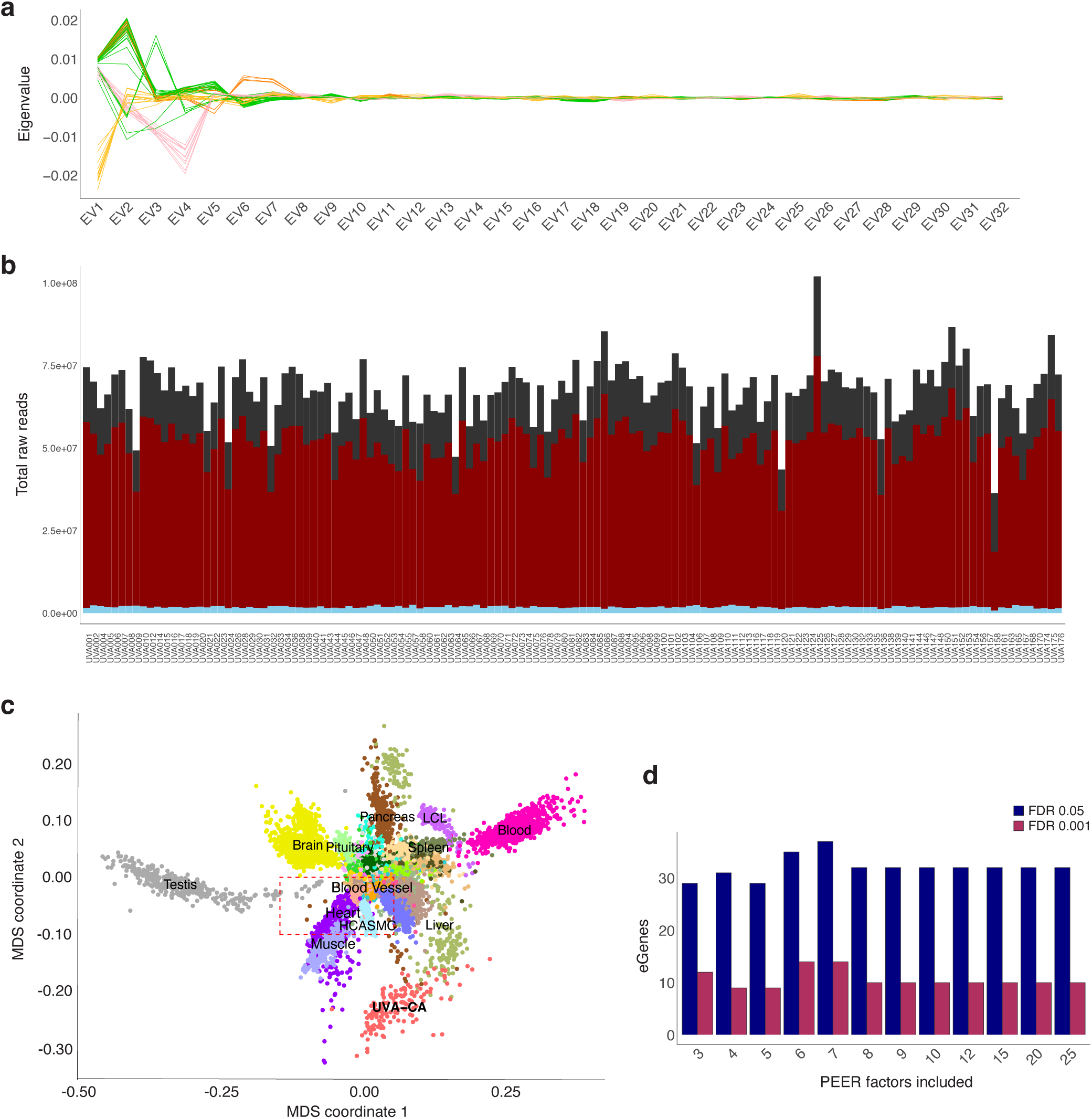
UVA sample genome-based ancestry and transcriptome-based expression values. (a) Line plot displays the values (y-axis) of 32 Eigenvectors (x-axis) calculated using the 1000G phase 3 reference panel, with each individual in our study population represented by a solid line. Colors correspond to Gencove-assigned majority continental ancestry: pink = Amerindigenous; blue = European; green = South Asian; orange = East Asian; yellow = African. (b) Cumulative raw read counts (y-axis) for lncRNA (light blue), protein-coding (red), and other (gray) genes annotated to GENCODE v32 for individuals considered for eQTL analyses (x-axis). (c) Multidimensional scaling (MDS) plot shows UVA transcriptomes clustered seprately from other GTEx tissue transcriptomes. Red dashed box includes overlapping tissue types of heart, muscle, and blood vessel and nearby HCAS<. (d) Bar plot represents the number (y-axis) of significant eQTLs identified among common variants within 500kb or 400 randomly selected chr17 genes adjusted for an increasing number of PEER factors included in the crude model (x-axis). FDRs of 5% and 0.1% are shown in navy blue and maroon, respectively.

**Fig S2.**
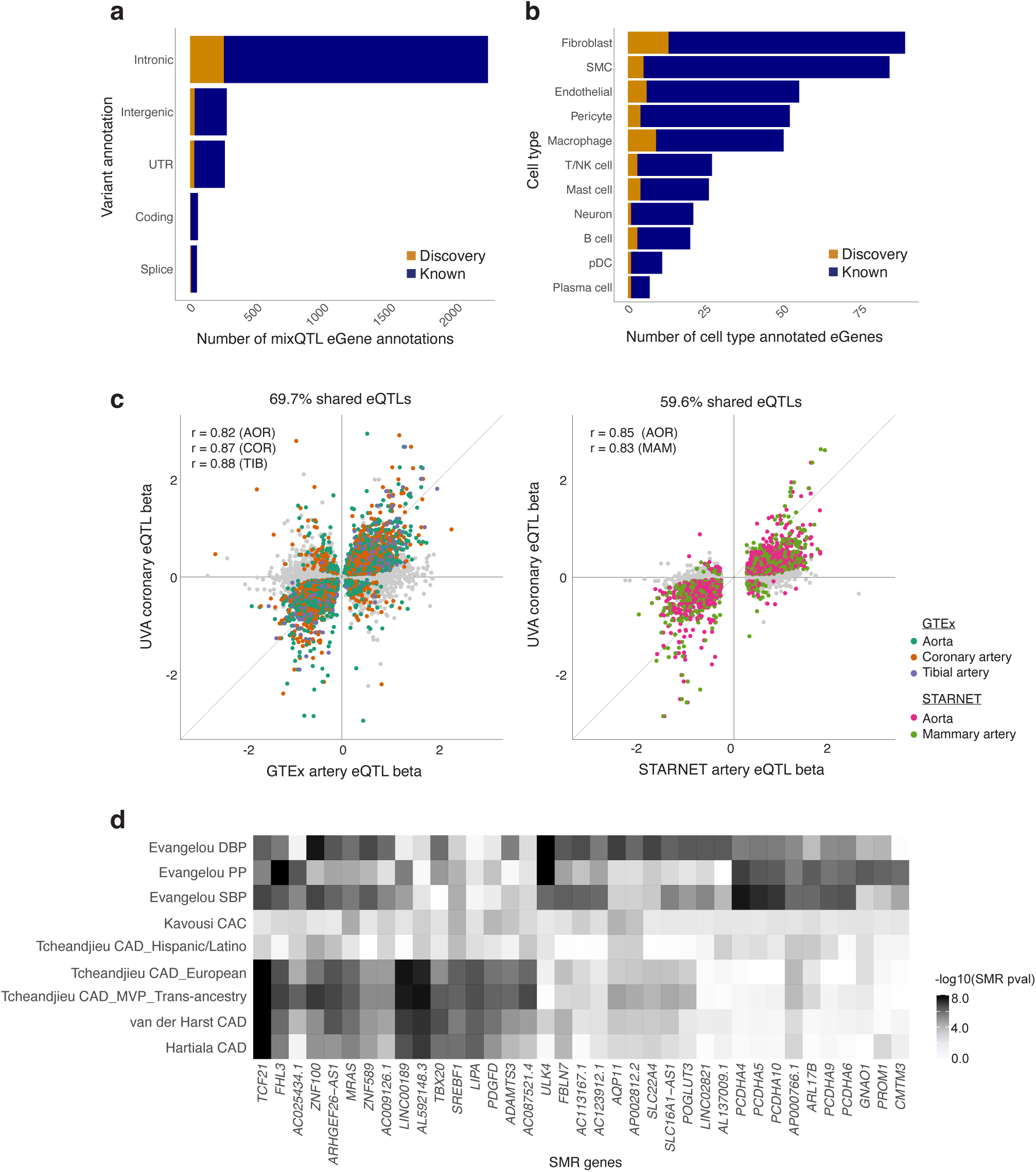
Annotation of mixQTL eGenes and shared effects in GTEx and STARNET artery tissues. (a) SNPEff eQTL-eGene annotations for lead eQTLs. Navy blue and orange represent numbers of reported and discovery eGenes, respectively. (c) Direction of effect for genes in which the UVA lead eQTL was significant (pBH<0.05) in the respective tissue. Pearson correlation coefficients (r) shown for overlapping significant UVA coronary eQTL detected in GTEx or STARNET eQTL with tissue indicated in parentheses. GTEx AOR: aorta (turquoise); COR: coronary artery (orange); TIB: tibial artery (purple); and STARNET AOR (pink); MAM: mammary artery (green). (d) Summarized Mendelian Randomization for GWAS and eGene associations: each column represents the -log10(SMR p-value) of the study (first author last name) and relevant GWAS trait, with intensity of shading corresponding to a more significant p-value. Each row represents one gene with SMR p<2.43E-7 (Bonferroni corrected for 2,057 tested genes) for at least one GWAS.

**Fig S3.**
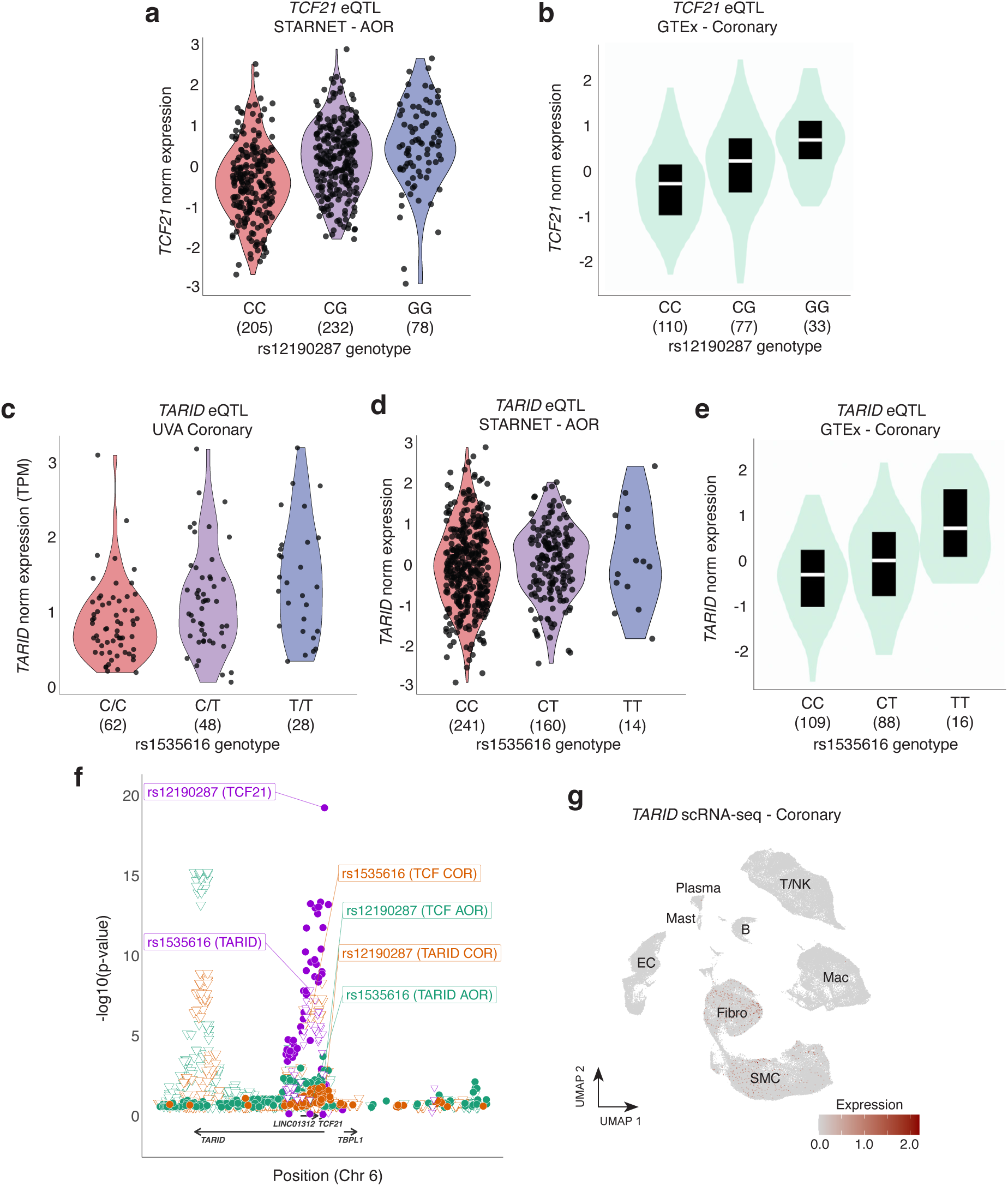
GWAS-eQTL colocalization eGenes in Europeans and supporting evidence for *TCF21/TARID* association. (a,b) Violin plots for normalized expression of *TCF21* (y-axis) in STARNET aortic root (AOR) tissue (a) or GTEx coronary artery tissue (b) shown by genotype (x-axis) for lead UVA eQTL rs12190287. (c-e) Violin plots for expression of *TARID* lncRNA (y-axis) in UVA coronary artery (c), STARNET AOR (d), or GTEx coronary artery tissue (e) shown by genotype (x-axis) for lead UVA eQTL rs1535616. (f) Associations of variants with gene expression for UVA coronary (purple), GTEx AOR (green), and GTEx COR (orange) for *TCF21* (circle) and *TARID* (triangle). Position and direction of gene coding regions shown below x-axis. (g) UMAP plot showing relative expression of *TARID* in single-cell RNA sequencing data from human artery reference dataset (Verdezoto Mosquera, et al, Biorxiv 2022). Each point represents a single cell; intensity of red color corresponds to higher relative expression of *TARID*. Broad cell type clusters are labeled. SMC: smooth muscle cells; EC: Endothelial cells, Fibro: Fibroblasts; Mac: Macrophage; B: B-cells; T/NK: T-cells and Natural Killer cells.

**Fig S4.**
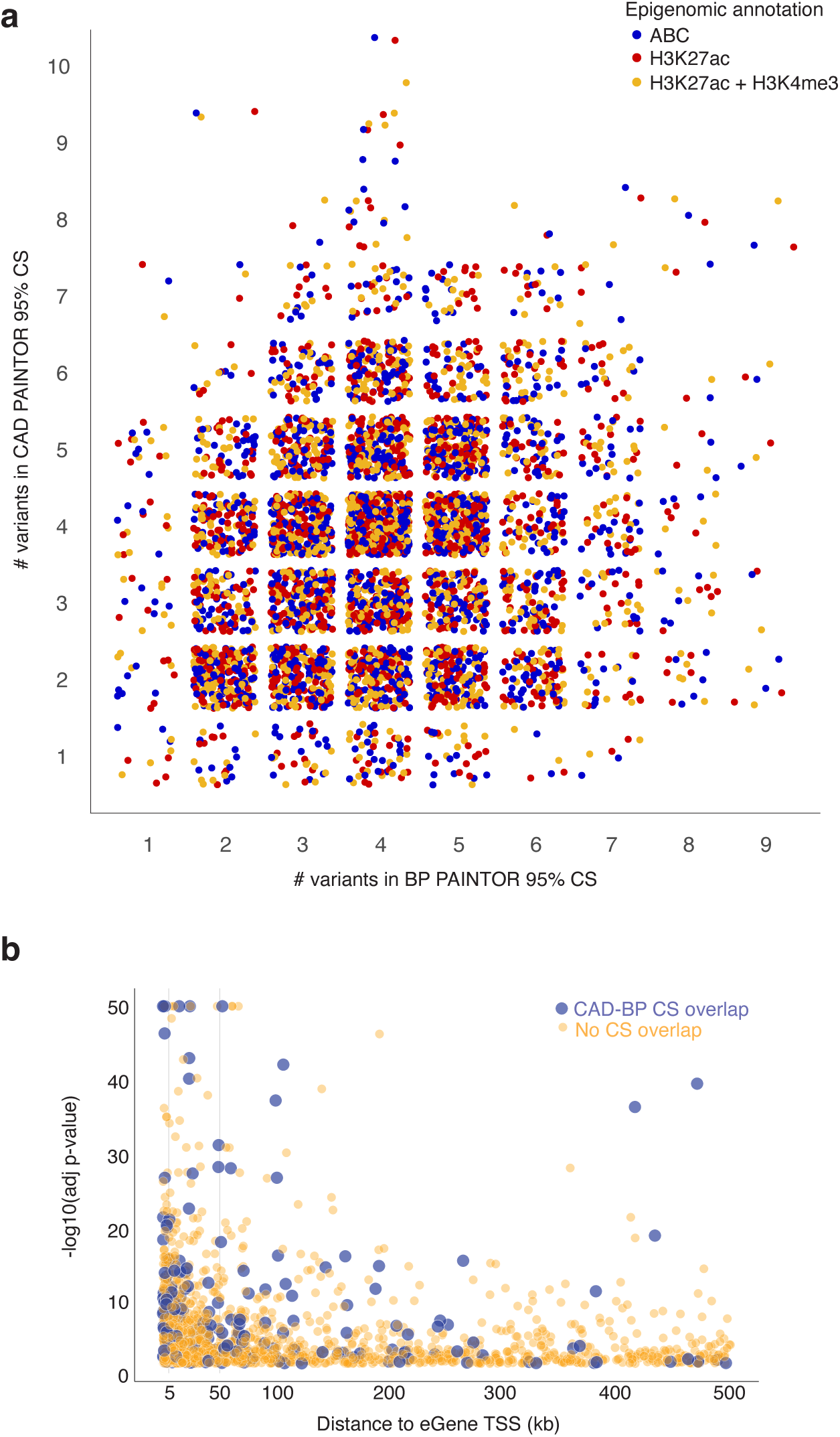
Fine-mapped credible set size and distance to TSS for CAD and BP traint GWAS. (a) Each point represents the number of variants in PAINTOR 95% credible set (CS) incorporating blood pressure trait GWAS data (x-axis) or CAD GWAS data (y-axis) for one UVA coronary artery eGene. All eGenes were fine-mapped using both GWAS datasets as well as annotation to ENCODE coronary artery activity-by-contact (ABC) chromatin contacts (blue), H3K27 acetylation (red), or H3K27 acetylation and H3K4 tri-methylation (yellow). For eGenes with no difference in credible set size only ABC annotation is shown. (b) Scatterplot shows significance of lead eQTLs (adjusted -log10(p-value), y-axis) versus distance from the corresponding eGene transcription start site (in kilobases, x-axis). Large blue points represent lead eQTLs for which at least one variant overlapped in the paintor CAD and BP GWAS-annotated credible sets; small orange points represent lead eQTLs for which no variants overlapped between the two credible sets for that eGene.

**Fig S5.**
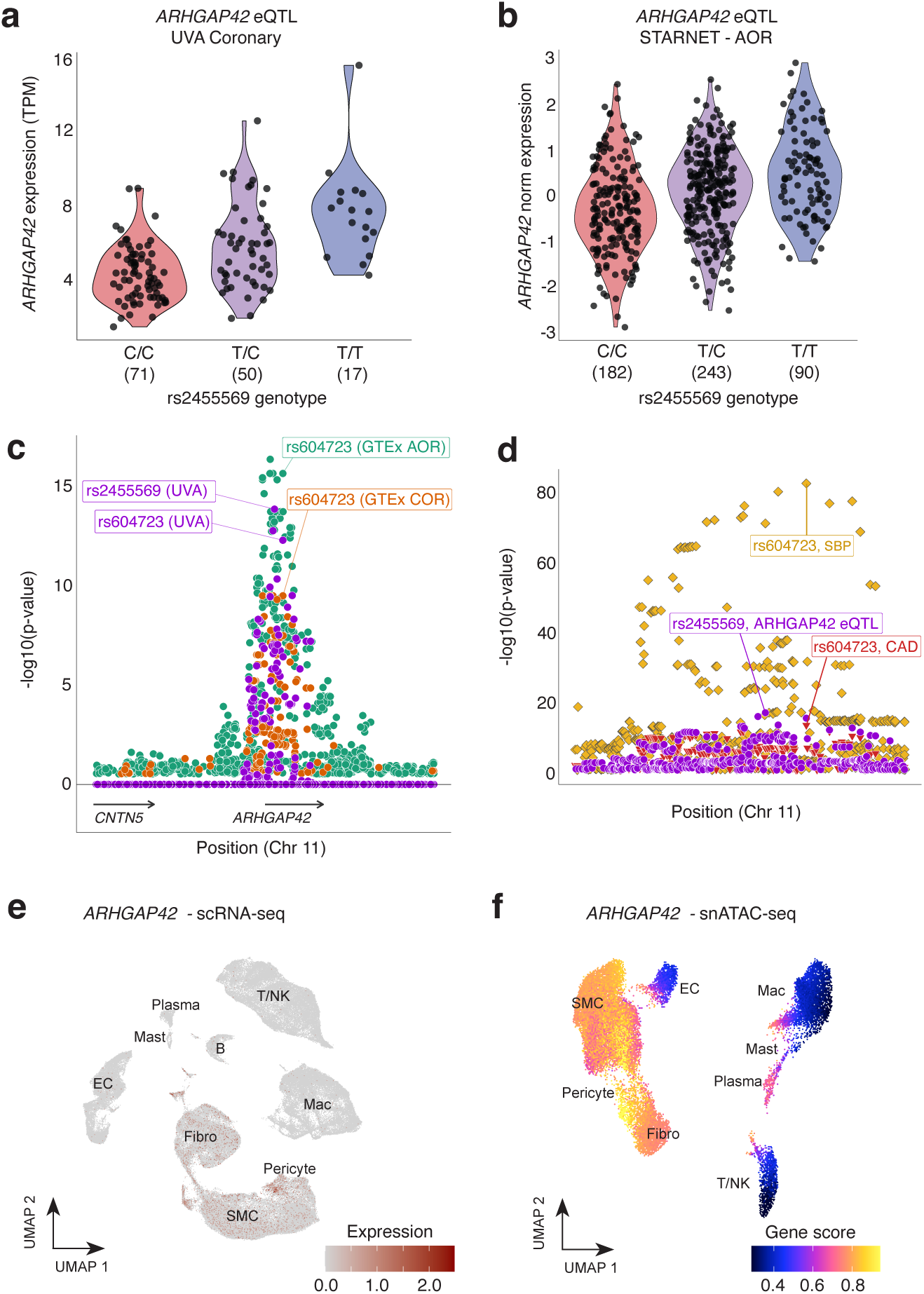
*ARHGAP42* eQTL and GWAS associations and cell type expression in human artery. (a,b) Violin plots for normalized expression of *ARHGAP42* (y-axis) in UVA coronary artery tissue (a) and STARNET aortic root tissue (AOR) (b) shown by genotype (x-axis) for lead UVA eQTL rs2455569. (c) Associations of individual variants with gene expression are plotted for UVA coronary (purple), GTEx AOR (green), and GTEx COR (orange) for *ARHGAP42* (circle) and *CNTN5* (triangle). (d) Association of individual variants are plotted for coronary artery gene expression (purple) or GWAS traits (red [CAD] and gold [SBP]). Position of variants on chromosome 11 is shown on the x-axis, with position and direction of gene coding regions shown underneath the x-axis. Significance of each variant shown as the -log10(p-value) of the association with expression of the respective gene on the y-axis. UVA lead variant rs2455569 is labeled. (e) UMAP plot showing relative expression of *ARHGAP42* in single-cell RNA sequencing data from human artery atherosclerosis reference dataset (Verdezoto Mosquera, et al, bioRxiv 2022). Each point represents a single cell; intensity of red color corresponds to higher relative expression of *ARHGAP42*. General cell type clusters are labeled accordingly. (f) UMAP plot showing imputed gene score activity at *ARHGAP42* calculated from a single-nucleus chromatin accessibility sequencing dataset in human coronary artery (Turner AW et al, Nat Genet 2022). SMC: smooth muscle cells; EC: Endothelial cells, Fibro: Fibroblasts; Mac: Macrophage; B: B-cells; T/NK: T-cells and Natural Killer cells.

**Fig S6.**
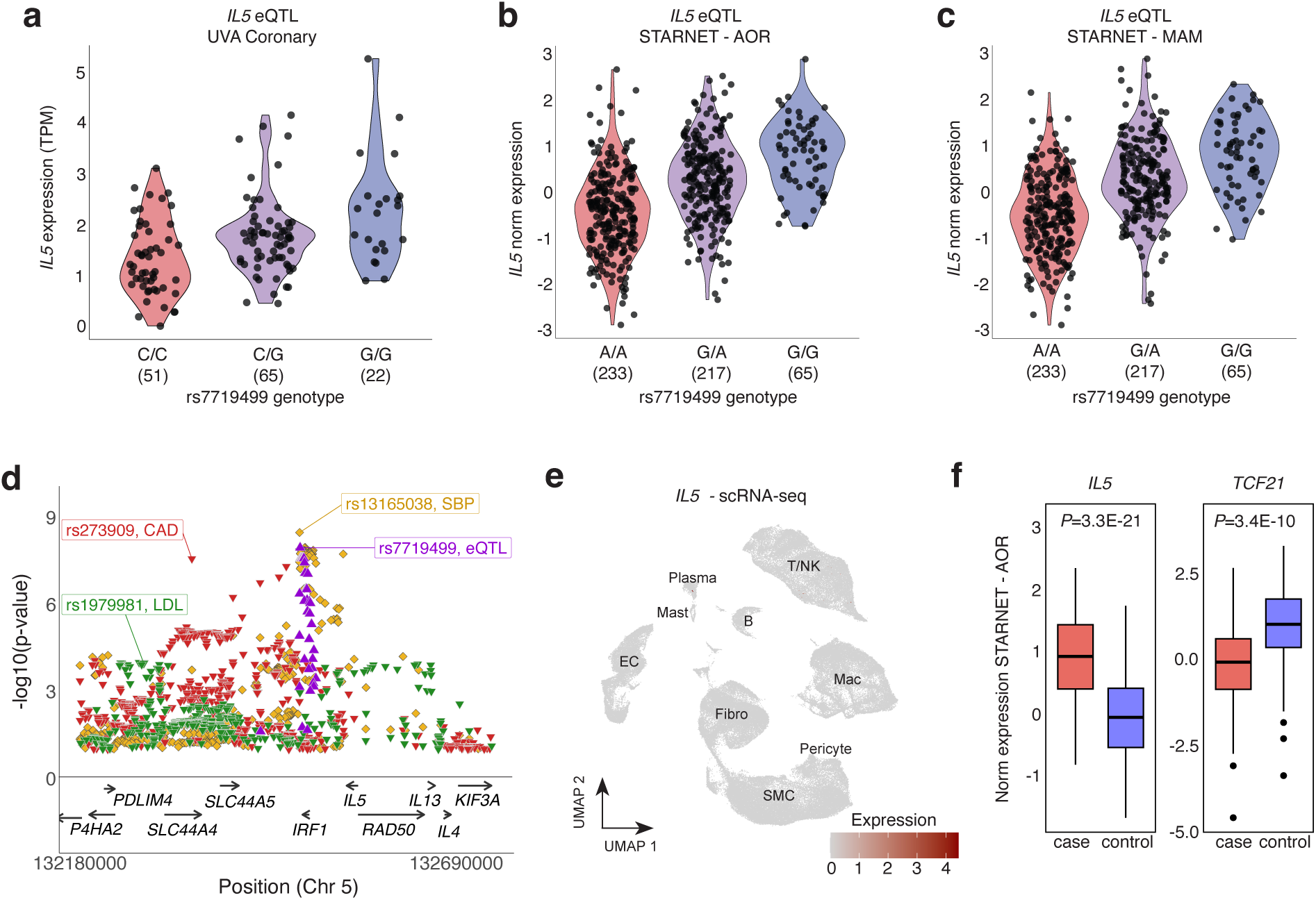
*IL5* eQTL associations, cell type and differential expression in human artery. (a-c) Violin plots for normalized expression of *IL5* (y-axis) in UVA coronary artery tissue (a), STARNET aortic root tissue (AOR) (b), and STARNET mammary artery tissue (c) shown by genotype (x-axis) for lead UVA eQTL rs7719499. (d) Association of individual variants are plotted for coronary artery gene expression (purple) or GWAS traits (red [CAD], gold [SBP], and green [LDL]). Position of variants on chromosome 5 is shown on the x-axis, with position and direction of gene coding regions shown underneath the x-axis. Significance of each variant shown as the -log10(p-value) of the association with expression of the respective gene on the y-axis. UVA lead variant rs7719499 is labeled. (e) UMAP plot showing relative expression of *IL5* in single-cell RNA sequencing data from human artery atherosclerosis reference dataset (Verdezoto Mosquera, et al, bioRxiv 2022). Each point represents a single cell; intensity of red color corresponds to higher relative expression of *ARHGAP42*. General cell type clusters are labeled accordingly. SMC: smooth muscle cells; EC: Endothelial cells, Fibro: Fibroblasts; Mac: Macrophage; B: B-cells; T/NK: T-cells and Natural Killer cells. (f) Normalized expression of *IL5* and *TCF21* in STARNET AOR tissues stratified by cases (individuals with coronary artery disease) or controls (individuals without coronary artery disease). Boxes represent upper and lower quartiles, line represents median, whiskers as 1.5 * IQR, and outliers as individual points. P-values were determined from DEseq2 and adjusted for metabolic phenotypes, drug treatments and principal components of ancestry (Koplev S, et al, Nat CVR 2022).

**Fig S7.**
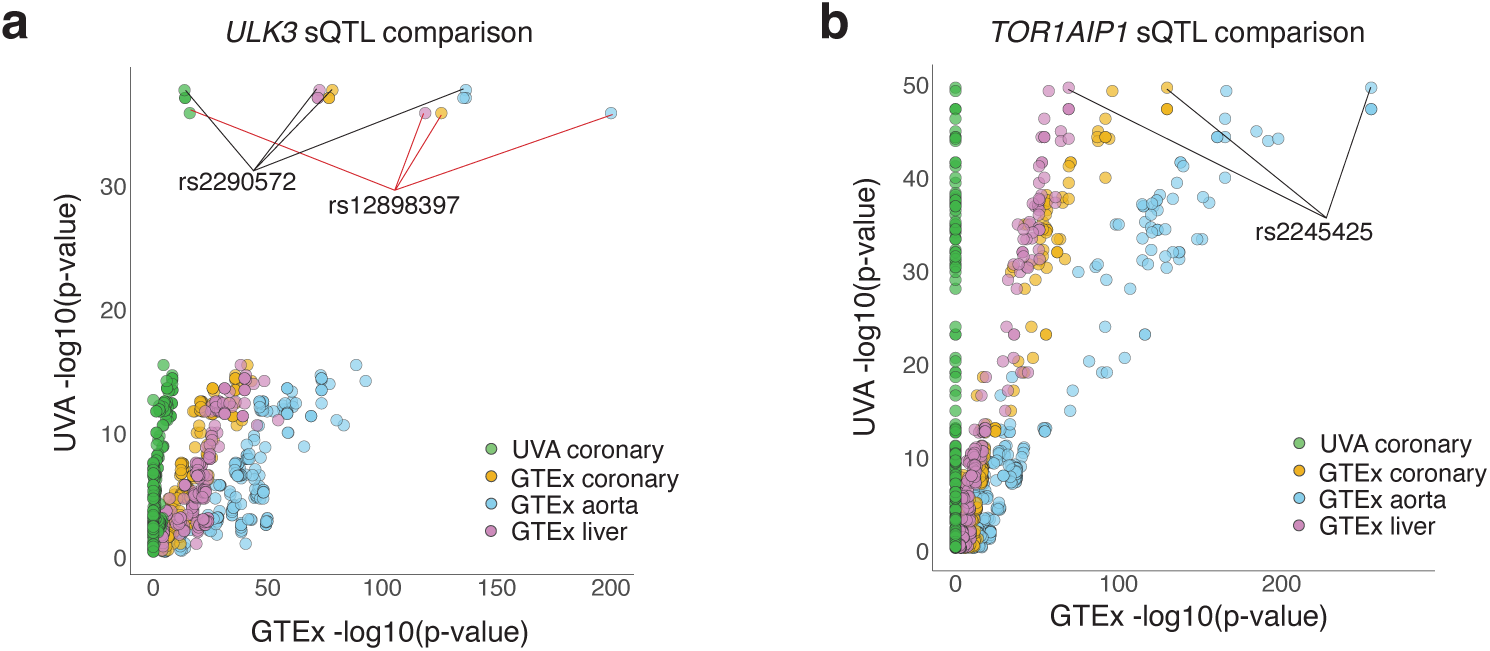
*ULK3* and *TOR1AIP1* sQTL association overlaps with GTEx tissues. (a) Scatterplot shows variants with UVA coronary sQTL significance (-log10(p-value) of association with chr15:74837435: 74837757 at *ULK3*, y-axis) compared to significance of the same variant in UVA eQTL for the same gene (green) and GTEx aorta (blue), coronary (orange), and liver (violet) tissues. All p-values are adjusted for the number of variants tested for that gene in the corresponding tissue. Lead UVA and GTEx QTLs (rs2290572 and rs12898397, respectively) are labeled. (b) Scatterplot shows variants with UVA coronary sQTL significance (-log10(p-value) of association with chr1:179884769:179889313 at *TOR1AIP1*, y-axis) compared to significance of the same variant in UVA eQTL for the same gene (green) and GTEx aorta (blue), coronary (orange), and liver (violet) tissues. All p-values are adjusted for the number of variants tested for that gene in the corresponding tissue. Lead UVA sQTL rs2245425 is labeled.

